# Social isolation and cardiovascular disease: mediation by health behaviours and metabolic risk factors in a 22-year survey follow-up

**DOI:** 10.1101/2025.08.18.25333293

**Authors:** Eero A.A. Teppo, Antti J. Etholén, Jaakko J. Harkko, Johannes Boch, Peter Speyer, Luka E.J. Vähäsarja, Jo I. Boufford, Pekka Jousilahti, Olli K. Pietiläinen, Tea M. Lallukka

## Abstract

**Background:** Social isolation is associated with cardiovascular disease (CVD). Few studies have explored whether this is mediated by health behaviours or metabolic risk factors in a longitudinal design.

**Methods:** Employees of the City of Helsinki (Finland) who turned 40, 45, 50, 55, or 60 years old were invited to participate in a Phase 1 survey in 2000–2 with four follow-ups up to 2022. The five phases were divided into three observation periods with three time points. Using a Bayesian predictive approach with logistic regressions, we estimated the effect of social isolation (living alone, meeting friends/relatives, social support, marital status) at the first time point on the risk of CVD in the third time point, with and without mediation by five health behaviours and four metabolic risk factors reported at the second time point.

**Results:** Among 5 403 participants (81% women, mean 49 years old in Phase 1) contributing 10 175 observations, a total of 846 (18%) participants reported a CVD in Phases 3–5. The total effect of social isolation on the risk of CVD was 1.01 (risk ratio; 90% credible interval 0.77–1.29), and the mediated effect through all mediators jointly was minimal along with other indirect pathways. Social isolation was associated with physical inactivity (prevalence ratio [PR] 1.41, 1.24–1.58) and low fruit and vegetable consumption (PR 1.47, 1.22–1.74).

**Conclusion:** Both the total and mediated effects of social isolation on CVD risk were modest. Social isolation should be considered when aiming to improve physical activity or nutrition in older populations.

## 1. Introduction

Social isolation, defined as an objective lack of interactions with others, is a growing public health concern shown to be associated with cardiovascular disease (CVD) risk and mortality. [1], [2] Many plausible mechanisms for such an effect have been proposed, from social factors (e.g. support for behaviour change) to psychobiological factors (e.g. chronic stress). [3], [4]

In meta-analyses of observational studies, different measures of social isolation have been associated with a higher incidence of stroke [5], coronary heart disease [6], CVDs [7], hospital readmission in heart failure [8], and CVD mortality [9]. Furthermore, randomised trials of cardiac rehabilitation [10] have suggested that social interventions could reduce CVD mortality, although this remains uncertain. The evidence is most convincing for the most severe outcomes like coronary and stroke deaths without hospital admission but less so for other CVD events [11]. In terms of potential mechanisms, social isolation has been associated with smoking, lower physical activity and fruit and vegetable consumption, and higher blood pressure and C-reactive protein levels, but also lower body mass index (BMI) and less alcohol consumption [12], [13]. Social support has been associated with better sleep [14] and adherence to treatments [15] and living alone associated with type 2 diabetes [16]. Isolated people also seek less outpatient care [17]. Physiological correlates have been explored even at the level of the blood proteome and metabolites and brain imaging [18], and while some candidates could be detected, such high-dimensional searches require strong replication.

The existing literature on the pathways between social isolation, key behavioural and metabolic factors, and morbidity holds several gaps [2]. Studies have often used cross-sectional designs for the exposure-mediator relationship, such as in an analysis of the UK biobank cohort [19], where adjusting for smoking, physical activity, and alcohol consumption reduced about one-third of the excess CVD mortality related to social isolation. Few analyses have more explicitly decomposed pathways from social isolation to CVD outcomes, which generally requires causal theory with strict assumptions [20]. In one analysis of the Third National Health and Nutrition Examination Survey (NHANES) cohort, a lifestyle index (smoking, physical activity, diet quality) could mediate about one-fifth of the total effect of social isolation on CVD-mortality [21]. Second, social isolation has been often measured with social network indices [22] that summarise different features of social isolation into a single number, which can hide relevant differences between the features. In one study, mostly frequent religious attendance and being married had clear associations when the score-components were analysed separately [23].

To better develop preventive interventions against cardiovascular disease, it is essential to clarify causal pathways and their relative importance. In mediation analysis, this involves distinguishing the total effect (the full impact of social isolation on CVDs, including all indirect and direct pathways), the indirect effect (the portion caused by intermediate factors, such as adverse health behaviours), and the direct effect (the portion caused by all remaining factors). Although these effects might not have a clear mechanistic interpretation even under a perfect trial design [24], a clear indirect effect would still be good evidence that the studied factors have some mediating role.

We had two linked research questions: 1) What are the direct, indirect, and total effects of social isolation on CVD through adverse health behaviours (physical inactivity, low fruit and vegetable consumption, smoking, heavy alcohol consumption, short sleep duration) and metabolic risk factors (obesity, diabetes, hypertension, hypercholesterolemia)? 2) Do these pathways vary among different aspects of social isolation (living alone, infrequent meetups, low social support, being single)?

## 2. Methods

### 2.1. Setting, design, and participants

The study population consisted of employees of the City of Helsinki, the capital of Finland. All employees turning 40, 45, 50, 55, or 60 years old during 2000–2002 were invited to participate in the Phase 1 survey, and respondents (67%) were sent follow-up surveys in Phases 2–5 in 2007 (response rate 83%), 2012 (79%), 2017 (82%), and 2022 (75%). [25]

We divided data from the five survey phases into three observation periods each with three time points: the first period included phases 1-2-3, the second period had phases 2-3-4, and the third period had phases 3-4-5 (illustrated in Supplementary Figure S1). One participant could thus contribute up to three observation periods and rows to the pooled dataset. This design was used to analyse the effect of social isolation in the first timepoint to the CVD risk in the third timepoint, through mediators in the second timepoint.

Observation periods were included if 1) they had no missing values in covariates (age, sex, socioeconomic position) or any mediators in the first time point, and if 2) they reported no history of CVD outcome in the second timepoint, that is, were at risk of CVD in the third time point. Out of 8 960 participants who responded to the Phase 1 survey, 5 403 (60%) contributed at least one observation to the analysis, giving a total of 10 175 observations out of which 3 066 (30%) had a missing CVD outcome. Out of the maximum possible 26 880 observations, first 2 134 (8%) were excluded for having past CVD, then 11 273 (46%) for missing CVD in the second time point, and finally 3 298 (24%) for missing covariates or mediators in the first time point. After the exclusions, exposures and mediators (second time point) had a few remaining missing values (less than 2.5%) which were imputed for analyses.

The study has been ethically approved by the health authorities of the City of Helsinki and the Ethical Committee of the Faculty of Medicine, University of Helsinki, Finland.

### 2.2 Exposure: Social isolation

Social isolation was defined as having a lower level of social interaction in four binary variables: living alone, low social support, being without a partner (marital status), and rarely meeting with friends and relatives. The main difference to commonly used measures like the Berkman-Syme index [26] is that we didn’t have access to repeated answers about participation in group activities and we have an item for social support. We also didn’t create a composite variable for social isolation but instead modelled them as separate variables without interactions. Thresholds for support and meetups were chosen so that approximately 20–30% were labeled as exposed to the factor.

Living alone (yes vs. no) was determined from the number of people reported living in the same household (partner, children, and other adults). The frequency of meetups with friends and relatives (less than twice per month vs. more) was measured by asking how many times participants met friends and relatives during the past four weeks and dichotomising the sum of the given categories’ means. Marital status was dichotomised by grouping participants into being with a partner (married or in a registered partnership) or without a partner (unmarried, divorced, separated, or widowed).

Social support (0–4 vs. 5+ points) was measured by asking participants ‘to evaluate your possibilities of getting help from your loved ones when you are in need of help or support’. This was asked for four different situations (stressed, depressed, practical help, or any possible situation) emphasizing ‘who can you really trust’, and six different roles (partner, friend, relative, colleague, neighbor, or other). Participants indicated whether they received support in each situation from a person in each role, with one point assigned per reported source of support. The total score, ranging from 0 to 24, was calculated by summing these points.

### 2.3. Mediators: Health behaviours and metabolic factors

Health behaviours measured included smoking (current regular vs. not), physical inactivity (less than 14 MET-hours per week vs. more) based on metabolic equivalents of tasks (18), low fruit and vegetable consumption (less than once a day vs. more), heavy alcohol consumption (7+ [women] or 14+ [men] units per week vs. less), and short sleep duration (6 or fewer hours by average on workdays vs. 7 or more). Metabolic CVD risk factors included obesity (BMI 30+ vs. less) and history of physician-diagnosed hypercholesterolemia, hypertension, and diabetes (yes vs. no). The measurement of these variables has been described in detail previously [27]. We also constructed three composite variables for having two or more adverse health behaviours, having two or more metabolic risk factors, and having four or more total risk factors.

### 2.4. Outcome: Cardiovascular disease

Measurement of CVD outcomes depends slightly on the survey phase and hence observations. In Phases 2–3 we had access to answers about the history of physician-diagnosed ‘angina pectoris’ or ‘heart attack’ as well as the Rose questionnaire, which is predictive of angina pectoris [28], [29]. In Phases 4–5, we had access to answers about the history of ‘coronary heart disease or other cardiovascular disease’ as well as the Rose questionnaire. Observations that had one or more positive items were classified with CVD; observations with negative reports on all items were classified as surviving the follow-up without CVD; and the rest were classified as lost to follow-up.

### 2.5. Covariates

Socioeconomic position was measured by occupational class (manual, routine non-manual, semi-professional, or managerial/professional), based on the job title acquired from the employer’s register for those who gave consent for register linkage and from the surveys for the rest. Analyses were also adjusted for age, sex (female vs. male), and observation period (1/2/3). When modelling mediators (second time point), analyses were additionally adjusted for their values in the first time point. Adjusting for baseline mediators is an effective way to reduce confounding in mediation analyses [30]. We didn’t add other factors as they are either rare, not available, or lack clear reasons. For example, adding comorbidities hasn’t previously changed estimates [12], [21].

### 2.6. Mediation and total effect analysis

We estimated total, direct, and indirect effects using a Bayesian counterfactual predictive (potential outcomes) approach, which corresponds to the Bayesian analog of parametric g-formula (non-iterative conditional expectation) estimator given necessary causal assumptions [31], [32], [33]. This is a general and intuitive framework which involves 1) fitting probabilistic parametric models conditioning on assumed biasing paths, 2) predicting outcomes under counterfactual interventions of interest, and 3) comparing the predictions to compute effect measures of interest.

We defined two models, one for the total effect of the four exposures without mediation, and one for the (natural) direct and indirect effects of the four exposures through one or more mediators. With both models, we compared predicted risks (CVD) and prevalences (mediators) for five pairs of counterfactual interventions: The effect of social isolation was defined as the ratio of the risk/prevalence had everyone received all four exposures and the risk/prevalence had no-one been exposed to any of the factors (and had no-one been lost to follow up in both). The effect of each exposure separately was defined similarly except for the other three exposures which are set to their observed values (no intervention).

We ran the total effect analysis separately for the CVD outcome and each mediator as outcome (Table 2) and the mediation analysis for each mediator separately but also for all metabolic factors jointly, all behavioural factors jointly, and all nine mediators jointly (Supplementary Table S2).

Specifically, we assumed the causal structure presented in Figure 1 and used logistic regression models. The log-odds of mediators were modelled as linear functions of the observation period, baseline value of the mediators, and exposures and covariates including a squared term for age. The log-odds of the CVD outcome was modelled similarly, adding the mediators (second time point) as predictors. Parameters had weakly informative Gaussian priors: intercept priors had a standard deviation (SD) of 5 (0.7–99.3% probability) and all coefficients an SD of 2 (odds ratio [OR] 0.14–7.40) as all prior means were set to zero (50% and OR 1). Parameter posteriors were inferred using a Hamiltonian Monte Carlo (HMC) sampler [34]. Predicted probabilities were simulated for all observations, including the ones with missing outcomes, and the effect posteriors were summarised using means and, for better stability, 90% credible intervals. All analyses were implemented in Stan [34] and R version 4.4.1 [35]; the script can be seen in the supplementary file.

**Figure 1.**
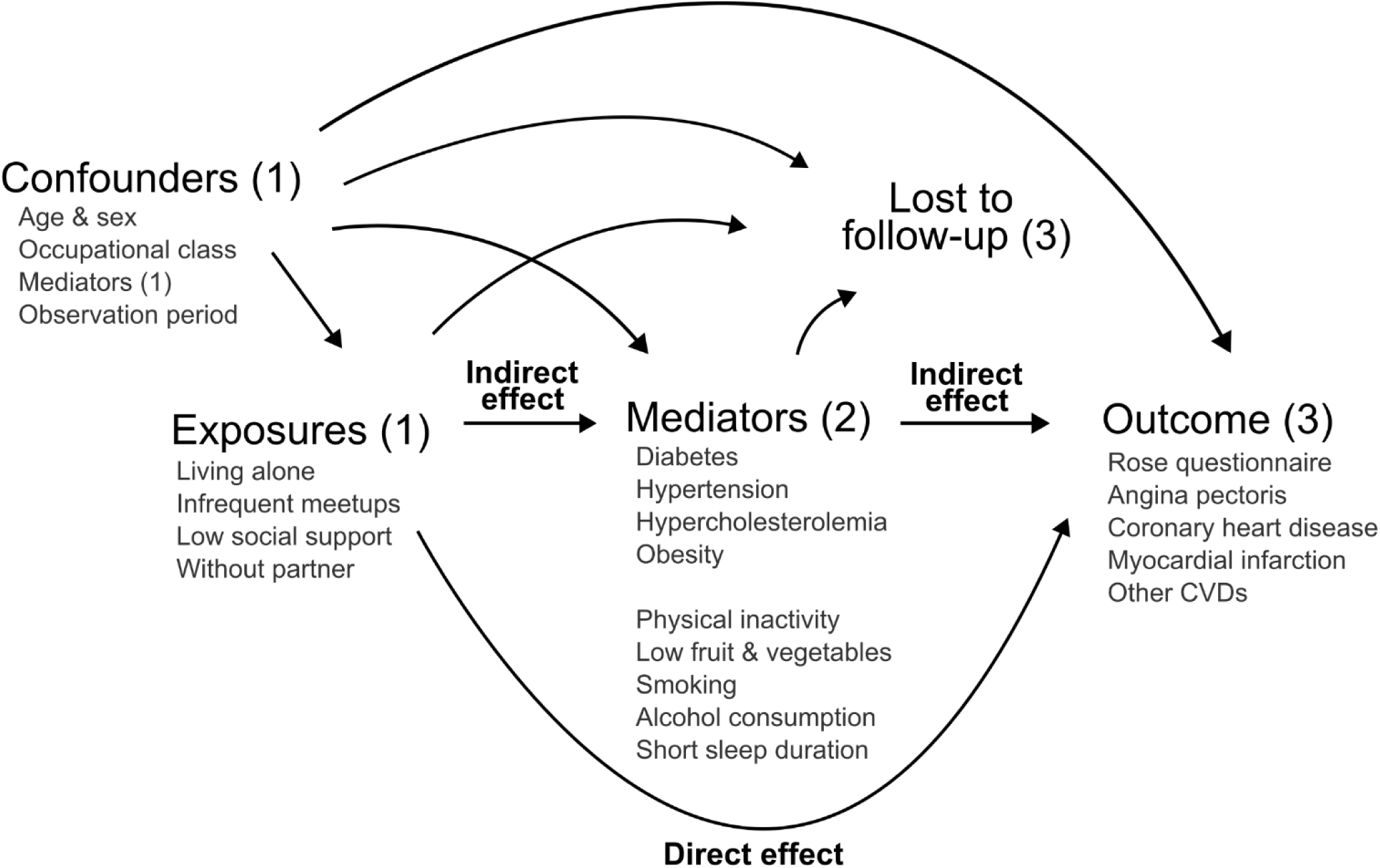
Causal assumptions in the mediation analyses estimating natural indirect and direct effects of the social isolation exposures on the cardiovascular disease (CVD) outcome. Numbers refer to time points of observation periods: The first observation period started in 2000–2 (1) with the midpoint (2) in 2007 and last point (3) in 2012. Similarly, for participants still at risk of outcome, the third observation period started in 2012 (1) with the midpoint for mediators in 2017 and the last time point for outcomes (3) in 2022. Alt text: A diagram describing the assumed causal relationships between study variables and loss to follow-up.

The remaining missing values in exposures and mediators (less than 2.5%) were single-imputed using logistic regression models based on baseline covariates and exposures, following the dependencies assumed in Figure 1.

### 2.6. Registry validation analysis

We assessed measurement error in self-reported diagnoses by validating the survey-based information on angina pectoris, myocardial infarction, cerebrovascular disease, claudication, as well as diabetes, hypertension, and hypercholesterolemia against national healthcare and prescription administrative register sources. (Supplement) Among the cohort participants who consented for register linkage (74–77% by survey), the reliability of hypertension and diabetes answers were over 90%, hypercholesterolemia over 80%, and Phase 2 heart attacks 87% and angina pectoris 52%. Survey answers were predictive of register diagnoses, with AUC values (area under the receiver operating characteristic curve) over 0.9, except for angina pectoris at 0.87. Mismatches were mostly false positives compared to registries.

## 3. Results

### 3.1. Descriptive results

Among the 5 403 included participants (81% women) with a mean age of 49 years in Phase 1, a total of 846 (18%) self-reported cardiovascular disease (CVD) in Phases 3–5, as 2 133 participants (40%) were lost to follow up (Supplementary Table S1).

A total of 1 222 participants (23%) reported living alone in Phase 1, of whom 22.0% reported CVD (vs. 18.9%; RR 1.16). (Supplementary Table S1) For other exposures, 35% reported a social support score of 0–4/24 (CVD 20.3% vs. 19.2%; RR 1.04), about 29% were single (CVD 20.2% vs. 19.4%; RR 1.04), and 17% reported meeting friends or relatives less than twice per month (CVD 20.4% vs. 19.6%; RR 1.04).

The prevalence of social isolation, defined as having all four exposures, was 1.2% in Phase 1 (up to 2.4% in Phase 5).

In Phase 2, 21% reported new hypercholesterolemia and 15% new hypertension while 8% changed obesity status and 3% reported new diabetes. The proportion of participants reporting a change in health behaviours between Phase 1 and 2 ranged from 8% for smoking to 23% for physical activity. (Supplementary Table S1)

More isolated participants tended to have a worse risk factor profile in Phase 1 (Table 1), but differences in background variables by social exposures were mostly small (Supplementary Table S1). The group living alone had slightly older participants, more men, and more participants who worked in managerial roles and tended to have a worse metabolic and behavioural profile in Phase 1. Other social exposures followed this pattern, with some exceptions such as singles including more women.

**Table 1.** Summaries (count [%] or mean) of the studied variables by composite social isolation and study inclusion in the Phase 1 survey. Data consisted of 8 960 employees of the City of Helsinki, Finland, who responded to a Phase 1 survey in 2000-2 when they were 40–60 years old.

| Variable | Summary | Social isolation* |  |  |  |  |
| --- | --- | --- | --- | --- | --- | --- |
|  |  | Included | High (3-4) | Moderate (1-2) | Low (0) | Excluded |
|  | N | 5403 | 478 | 2672 | 1935 | 3564 |
| **Background** |  |  |  |  |  |  |
| Age | Years (mean) | 48.8 | 50.6 | 49.2 | 47.9 | 50.2 |
| Occupational class | Manual [N (%)] | 1737 (32.1**) | 148 (31.0) | 913 (34.2) | 584 (30.2) | 1236 (36.6) |
|  | Routine non-manual | 1164 (21.5) | 78 (16.3) | 552 (20.7) | 468 (24.2) | 551 (16.3) |
|  | Semi-professional | 1767 (32.7) | 147 (30.8) | 818 (30.6) | 702 (36.3) | 833 (24.7) |
|  | Manager / Professional | 735 (13.6) | 105 (22.0) | 389 (14.6) | 181 (9.4) | 757 (22.4) |
| Sex | Female | 4365 (80.8) | 378 (79.1) | 2137 (80.0) | 1587 (82.0) | 2803 (78.8) |
| **Exposures: Social isolation** |  |  |  |  |  |  |
| Meetups with friends/relatives | < 2 meetups / month | 850 (16.6) | 171 (35.8) | 675 (25.3) | 0 (0.0) | 581 (17.9) |
| Living alone | Yes | 1222 (22.7) | 429 (89.7) | 727 (27.2) | 0 (0.0) | 960 (27.3) |
| Marital status | Single | 1569 (29.2) | 452 (94.6) | 1019 (38.1) | 0 (0.0) | 1159 (32.8) |
| Social support | Score 0–4 | 1886 (35.1) | 445 (93.1) | 1328 (49.7) | 0 (0.0) | 1585 (45.2) |
| **Mediators: Metabolic factors** |  |  |  |  |  |  |
| Diabetes | Has history | 112 (2.2) | 16 (3.5) | 62 (2.5) | 26 (1.4) | 125 (4.5) |
| Hypercholesterolemia | Has history | 1039 (19.3) | 99 (20.7) | 517 (19.4) | 361 (18.7) | 846 (24.2) |
| Hypertension | Has history | 1067 (19.8) | 96 (20.1) | 530 (19.9) | 377 (19.6) | 964 (27.7) |
| Obesity | BMI 30+ | 677 (12.6) | 73 (15.4) | 352 (13.3) | 216 (11.2) | 598 (17.2) |
| **Mediators: Health behaviours** |  |  |  |  |  |  |
| Alcohol consumption | > 7/14 units per week (women / men) | 860 (16.3) | 83 (17.8) | 411 (15.7) | 319 (16.7) | 551 (16.8) |
|  |  | Included | High (3-4) | Moderate (1-2) | Low (0) | Excluded |
| Fruit & vegetables | < 1 serving / day | 641 (11.9) | 95 (20.0) | 333 (12.5) | 174 (9.0) | 540 (15.5) |
| Physical activity | < 14 MET hours / week | 1235 (23.0) | 129 (27.2) | 648 (24.4) | 387 (20.1) | 1002 (28.6) |
| Sleep duration | < 7 hours / workday | 1196 (22.4) | 136 (28.7) | 599 (22.7) | 365 (19.0) | 906 (26.3) |
| Smoking | Current regular | 1123 (20.9) | 120 (25.3) | 565 (21.2) | 367 (19.1) | 993 (28.4) |

\* The number of exposures (living alone, rare meetups, low support, being single) in Phase 1.
\*\* Missing values were excluded when calculating percentages.
Abbreviations: BMI, body mass index; MET, metabolic equivalents of task.

### 3.2. Total effect results

The effect of social isolation (all vs. none) on CVD risk was RR 1.01 (90% credible interval 0.77–1.29). Estimates for each exposure separately were RR 1.12 (0.94–1.33) for living alone, RR 0.95 (0.78–1.13) for infrequent meetups, RR 0.96 for low perceived support (0.84–1.08), and RR 1.00 (0.84–1.18) for being without a partner. (Table 2)

**Table 2.** Estimated total effects of social isolation exposures at baseline on potential mediators (second time point) and cardiovascular disease (third time point). The analyses adjusted for age, sex, and occupational class, as well as for baseline value of each mediator and observation period. Effects are presented as risk/prevalence ratios and their 90% credible intervals. The data consisted of 10 175 observations from 5 403 participants in the Helsinki Health Study of employees of the City of Helsinki, Finland.

| Outcome | Exposure |  |  |  |  |
| --- | --- | --- | --- | --- | --- |
|  | Social isolation<br>(all vs. none) | Living alone | Rare meetups<br>(< 2/month) | Low<br>social support<br>(0-4 points) | Single<br>(marital status) |
| Cardiovascular disease | 1.01 (0.77-1.29) | 1.12 (0.94-1.33) | 0.95 (0.78-1.13) | 0.96 (0.84-1.08) | 1.00 (0.84-1.18) |
| **Metabolic factors** |  |  |  |  |  |
| Hypertension | 1.04 (0.97-1.12) | 1.02 (0.97-1.07) | 1.02 (0.97-1.07) | 1.03 (0.99-1.07) | 0.98 (0.93-1.03) |
| Diabetes | 1.04 (0.86-1.25) | 1.10 (0.96-1.25) | 0.97 (0.85-1.10) | 0.97 (0.89-1.07) | 1.01 (0.89-1.15) |
| Hypercholesterolemia | 1.04 (0.96-1.13) | 1.04 (0.98-1.09) | 1.05 (0.99-1.10) | 1.01 (0.97-1.05) | 0.95 (0.90-1.00) |
| Obesity | 1.08 (0.97-1.19) | 1.09 (1.01-1.17) | 1.02 (0.95-1.09) | 0.99 (0.94-1.04) | 0.98 (0.92-1.05) |
| **Health behaviours** |  |  |  |  |  |
| Smoking | 1.16 (1.05-1.27) | 1.03 (0.95-1.10) | 1.06 (0.98-1.13) | 1.01 (0.96-1.07) | 1.06 (0.99-1.13) |
| Heavy alcohol<br>consumption | 0.96 (0.85-1.08) | 0.91 (0.83-0.99) | 1.06 (0.98-1.15) | 0.98 (0.93-1.04) | 1.02 (0.94-1.10) |
| Short sleep duration | 1.12 (0.99-1.25) | 1.01 (0.93-1.09) | 1.05 (0.97-1.13) | 1.02 (0.96-1.08) | 1.04 (0.96-1.12) |
| Low fruit & vegetable<br>consumption | 1.47 (1.22-1.74) | 0.99 (0.86-1.13) | 1.20 (1.06-1.35) | 1.07 (0.97-1.17) | 1.16 (1.02-1.32) |
| Physical inactivity | 1.41 (1.24-1.58) | 0.97 (0.89-1.06) | 1.14 (1.05-1.23) | 1.10 (1.03-1.17) | 1.16 (1.07-1.26) |
| **Mediator composites** |  |  |  |  |  |
| 2+ metabolic factors | 1.04 (0.95-1.14) | 1.05 (0.98-1.12) | 0.99 (0.92-1.05) | 1.02 (0.98-1.07) | 0.98 (0.92-1.04) |
| 2+ behavioural factors | 1.27 (1.14-1.41) | 0.97 (0.89-1.05) | 1.10 (1.02-1.18) | 1.05 (0.99-1.11) | 1.14 (1.06-1.23) |
| 4+ risk factors | 1.21 (1.03-1.41) | 1.05 (0.94-1.17) | 1.04 (0.93-1.15) | 1.02 (0.94-1.11) | 1.09 (0.98-1.21) |

Social isolation, however, was associated with physical inactivity (prevalence ratio 1.41, 1.24–1.58) and low fruit and vegetable consumption (PR 1.47, 1.22–1.74) with the largest contributions coming from being single and infrequent meetups. This was also reflected in the association with having multiple behavioural risk factors (PR 1.27, 1.14– 1.41) and four or more risk factors (PR 1.21, 1.03–1.41). The overall pattern was that associations were minimal but more likely to be detrimental for health, with some exceptions such as living alone associated with less heavy alcohol consumption (PR 0.91, 0.82–0.99).

### 3.3. Indirect (mediated) and direct (remaining) effect results

Results for each mediator separately (Supplementary Table S2) confirmed that the indirect effects of social isolation on CVD risk were negligible, and the remaining direct effects were consistent with the total effect estimates, independent of which mediator was modelled. Results were the same for multiple mediators with an indirect effect of RR 1.03 (90% credible interval 1.00–1.05) for all mediators jointly, and RR 1.00 (0.99–1.2) for metabolic and RR 1.03 (1.01–1.05) for behavioural factors.

## 4. Discussion

### 4.1 Main findings

We studied the effect of social isolation on the risk of cardiovascular disease (CVD) and whether it was mediated by health behaviours or metabolic risk factors. We found that the effect of social isolation and its components on CVD risk was overall small or non-existent. Social isolation was associated with physical inactivity and low fruit and vegetable consumption, which was mostly explained by infrequent meetups and being without a partner, but overall associations between exposures and mediators were small.

### 4.2. Strengths & limitations

This study utilised longitudinal data spanning 22 years across five phases with relatively high participation across surveys, allowing us to observe changes in many potential mediators over 5–7 years and then new CVDs over five years after. We were able to capture many kinds of social interaction (with the notable exception of work relationships) combined with how supportive they were perceived to be, and then able to explore their relative contributions in total and mediated effects.

Risks to the validity of our results include potential confounding, selection bias, and measurement error typical of survey-based health studies, as well as model misspecification. Causal interpretations can be also invalid due to lack of consistency which informally means that the exposure data doesn’t match with well-defined counterfactual interventions.

First, even though our design and analysis should reduce various sources of confounding, conditional exchangeability within the chosen covariates cannot be strongly justified. The causal structure is not well-known. We didn’t adjust for prior exposure, and although social interaction is highly modifiable, our exposures aren’t generally consistent with realistic interventions (e.g. making everyone live alone in Phase 1). This reveals another potential source of bias, interference, as social factors can have complex network effects.

Second, non-response at each phase can induce selection bias. Associations between survey response and various socioeconomic and health variables were minor in the early phases [25] which may limit the risk of bias from the exclusions due to missing data. Loss to follow-up was relatively high, and the analyses assumed this to be avoidable non-response and random given the exposures, mediators, and covariates, but relevant factors could’ve been missed.

Third, significant measurement bias could arise from the survey answering process and undetected or misdiagnosed conditions. We validated the survey-based diagnoses against more objective registry data and the accuracy was acceptable for the studied variables which increased credence that false positives or other errors didn’t dominate the results. The use of binary variables also limits the impact of small errors in other variables.

### 4.3. Comparison with previous results

Previous studies have found social isolation to increase the risk of CVD outcomes [5], [6], [7], [8], [9]. Apart from different sources of error, a smaller association in our study could be explained by several factors: First, studies have analysed different questions, e.g., there are differences in defining and measuring social isolation and CVD outcomes.

The prevalence of severe isolation was low in our cohort which may have limited the ability to detect clear differences. Furthermore, our outcome was defined to capture cases broadly including symptoms indicative of coronary heart disease, as well as other diseases of the cardiovascular system. Second, context-specific factors, such as cultural, social, or healthcare differences between study populations, may influence the relationship between social isolation and CVD. Populations with stronger public safety nets are expected to be less affected by individual isolation. For instance, access to healthcare and other community support in our cohort could have buffered against certain effects. Especially emotional responses to different forms of isolation can be culture dependent. Our results on health behaviours and obesity as outcomes generally agree with previous studies [12], [13], although the associations are smaller, and we could only make confident statements about the increase in physical inactivity and low fruit and vegetable consumption in more isolated groups.

Finally, previous studies have suggested that health behaviours could explain a fraction of CVD harms of social isolation [19], [21]. In our cohort, neither health behaviours nor metabolic factors appeared to mediate effects on CVD, although the lack of a total effect made estimation difficult. One possible reason could be that our follow-up was not long and granular enough to capture effects of longer-term isolation, adverse behaviours, and metabolic risks. For example, hypercholesterolemia likely increases CVD risk by accumulating into arterial plaques over the lifespan [36], so analysing effects through new hypercholesterolemia diagnoses in older adults can miss much of the total effect. However, the benefit is that any indirect effects estimated are more likely to be true rather than a result of unknown confounding from the past.

### 4.4. Conclusion

While social isolation is associated with some differences in health behaviours, its impact on cardiovascular outcomes appears minimal in older adults in this context. Addressing social isolation should be considered when aiming to improve physical activity and nutrition in older populations. Public health efforts should continue to address modifiable risk factors and promote social connectedness, but additional research is needed to better understand the pathways linking social isolation to health outcomes and to identify other potential mediators or modifiers in different contexts.

## Data Availability

The Helsinki Health Study survey data cannot be shared due to strict data protection laws and regulations. More information is available at the project website (helsinki.fi/hhs), in our data protection statement (helsinki.fi/hhs/data-protection-statement), and from. Permission to the secondary use of health and social registry data is granted by the Finnish Social and Health Data Permit Authority, Findata.

## 5. Conflicts of interest

None.

## 6. Funding

This research was supported by the Novartis Foundation and the Research Council of Finland (Grant #330527).

## 7. Key points

- Social isolation in older adults was not strongly associated with an increased risk of cardiovascular disease 5-12 years later in life, neither directly nor through health behaviours or metabolic risk factors.
- Social isolation was associated with lower physical activity and low fruit and vegetable consumption, but these changes were not substantial enough to change CVD risk in this population.
- Differences in definitions, study populations, and healthcare systems, among other factors, may explain why prior studies found stronger associations, highlighting the need for further research on long-term effects and potential mediators.

## Notes

### Competing Interest Statement

The authors have declared no competing interest.

### Funding Statement

This study was supported by the Novartis Foundation and the Research Council of Finland (Grant #330527).

### Author Declarations

Ethics committee of the University of Helsinki Faculty of Medicine gave ethical approval for this work.

