## Supplementary information for "Social isolation and cardiovascular disease: mediation by health behaviours and metabolic risk factors in a 22-year survey follow-up"

**Supplementary information for "Social isolation and cardiovascular disease: mediation by health behaviors and metabolic risk factors in a 22-year survey follow-up"**

### 1. Register validation of survey-based diagnosis data

#### 1.1. Methods

We tested our survey-based measures of cardiovascular outcomes (heart attack, cerebrovascular disease, angina pectoris, claudication) and metabolic risk factors (hypertension, hypercholesterolemia, and diabetes) against register-based data, which included International Classification of Diseases (ICD-10) codes (1) and medications classified under the Anatomical Therapeutic Chemical (ATC) codes (2).

Consent rates for register linkage for each phase (Validation Table 1) were 74% in Phase 1, 76% in Phase 2, 76% in Phase 3, 76% in Phase 4, and 77% in Phase 5. Register data were obtained from the Social Insurance Institution of Finland (3), the Care Register for Health Care (inpatient care) (4), and the Register of Primary Health Care Visits (5). The final analytic sample for the validation study comprised 6603 individuals in Phase 1, 5573 individuals in Phase 2, 5197 individuals in Phase 3, 5206 individuals in Phase 4, and 4587 individuals in Phase 5.

The data were analyzed following national data protection laws in Kapseli, a secure operating environment for analyzing sensitive individual level data under the Act on the Secondary Use of Health and Social Data. (6) Participants consenting and non-consenting to register data linkage were broadly similar, although age, occupational class, and sickness absence were somewhat associated with consent. (7)

From the register of the Social Insurance Institution of Finland, we used data on reimbursed essential (primary) hypertension medications (ATC codes C02, C03, C07, C08, C09), pure hypercholesterolemia (C10AA, C10AB, C10AX), and diabetes (A10A, A10B) medication purchases from 2000 to 2022. (2) For diagnostic codes (ICD-10) from 1990–2022 in the Care Register for Health Care (inpatient care) and the Register of Primary Health Care Visits, we used angina pectoris (I20), myocardial infarction (I21), cerebrovascular disorder (I60–I69), intermittent claudication (I73.9), essential hypertension (I10), pure hypercholesterolemia (E78.0) and diabetes (E10, E11, E13, E14). (1) Events related to medication purchases or ICD diagnoses were considered up to the date of each respective phase's questionnaire submission, as the survey question asked, 'Has a doctor ever diagnosed you with any of the following conditions?'.

Data from all register sources were merged for each testing measurement to examine how the register data matched with self-reported data from surveys (Validation Table 2). First, the prevalence of register-based measurements compared to survey-based measurements was calculated (Validation Table 3). Next, age-adjusted log-binomial regression models were fitted with register-based measurements as the outcome and survey-based measurements as the predictor to calculate prevalence ratios (PRs) and their 95% confidence intervals (95% CIs). Finally, receiver operating characteristic (ROC) curve analyses were conducted to test the diagnostic performance of the survey data in relation to the register data. (8) In addition, the reliability of our survey-based measurements was tested between phases (Validation Table 4).

#### 1.2. Results

##### Validation Table 1: Register linkage consent counts and percentages by survey phase for all participants and survey-based diagnostic subgroups

Validation Table 1. Consents of register linkages among the participants of the Helsinki Health Study in 2000-2002 (Phase 1, age 40-60 years), 2007 (Phase 2), 2012 (Phase 3), 2017 (Phase 4) and 2022 (Phase 5, age 62–82 years).

| | Phase 1 | Phase 2 | Phase 3 | Phase 4 | Phase 5 |
|---|---|---|---|---|---|
| | % | % | % | % | % |
| **All participants (N)** | **8959** | **7332** | **6808** | **6831** | **5950** |
| No consent information | 8 | 8 | 8 | 7 | 7 |
| With consent | 74 | 76 | 76 | 76 | 77 |
| Without consent | 18 | 16 | 16 | 16 | 16 |
| **Metabolic risk factors** | | | | | |
| **Hypertension (N)** | **2031** | **2373** | **2584** | **2799** | **2999** |
| No consent information | 10 | 9 | 9 | 8 | 8 |
| With consent | 74 | 75 | 75 | 76 | 77 |
| Without consent | 16 | 16 | 16 | 16 | 15 |
| **Hypercholesterolemia (N)** | **1884** | **2527** | **2926** | **2333** | **NA** |
| No consent information | 9 | 8 | 8 | 7 | NA |
| With consent | 76 | 76 | 77 | 78 | NA |
| Without consent | 15 | 16 | 15 | 15 | NA |
| **Diabetes (N)** | **237** | **389** | **607** | **803** | **NA** |
| No consent information | 10 | 9 | 9 | 10 | NA |
| With consent | 76 | 77 | 73 | 74 | NA |
| Without consent | 14 | 14 | 17 | 16 | NA |
| **Cardiovascular outcome** | | | | | |
| **Angina Pectoris (N)** | **498** | **149** | **154** | **NA** | **NA** |
| No consent information | 11 | 9 | 8 | NA | NA |
| With consent | 70 | 77 | 72 | NA | NA |
| Without consent | 19 | 14 | 20 | NA | NA |
| **Heart attack (N)** | **60** | **86** | **110** | **NA** | **NA** |
| No consent information | 13 | 9 | 9 | NA | NA |
| With consent | 65 | 79 | 75 | NA | NA |
| Without consent | 22 | 12 | 16 | NA | NA |

| | | | | | |
|---|---|---|---|---|---|
| **Claudication (N)** | 21 | 33 | 33 | NA | NA |
| No consent information | 10 | 12 | 21 | NA | NA |
| With consent | 67 | 67 | 61 | NA | NA |
| Without consent | 24 | 21 | 18 | NA | NA |
| **Cerebrovascular disease (N)** | **113** | **144** | **198** | **NA** | **255** |
| No consent information | 12 | 10 | 11 | NA | 8 |
| With consent | 79 | 78 | 76 | NA | 79 |
| Without consent | 10 | 12 | 13 | NA | 13 |

**Validation Table 2: Prevalence of register-based diagnoses by survey-based diagnoses and survey phase**

Validation Table 2. The prevalence of register-based measurements compared to survey-based measurements among the participants of the Helsinki Health Study who consented for register linkage. NA = variable was not measured in the survey.

| Metabolic risk factors | Phase I | | Phase II | | Phase III | | Phase IV | | Phase V | |
|---|---|---|---|---|---|---|---|---|---|---|
| **Hypertension** | **Yes** | **No** | **Yes** | **No** | **Yes** | **No** | **Yes** | **No** | **Yes** | **No** |
| Participants (N) | 1505 | 5046 | 1790 | 3690 | 1948 | 3176 | 2117 | 2048 | 2304 | 1590 |
| By ICD-codes: I10 (%) | | | | | | | | | | |
| No | 94 | 100 | 89 | 100 | 55 | 99 | 28 | 96 | 19 | 94 |
| Yes | 5 | 0 | 11 | 0 | 45 | 1 | 72 | 4 | 81 | 6 |
| By ATC-codes: C02, C03, C07, C08, C09 (%) | | | | | | | | | | |
| No | 32 | 96 | 13 | 88 | 5 | 81 | 2 | 74 | 3 | 68 |
| Yes | 68 | 4 | 87 | 12 | 95 | 19 | 98 | 26 | 97 | 32 |
| **Hypercholesterolemia** | **Yes** | **No** | **Yes** | **No** | **Yes** | **No** | **Yes** | **No** | **Yes** | **No** |
| Participants (N) | 1439 | 5110 | 1928 | 3592 | 2264 | 2908 | 1821 | 2096 | 2044 | 1575 |
| By ICD-codes (%) | | | | | | | | | | |
| No | 100 | 100 | 100 | 100 | 87 | 99 | 67 | 94 | 57 | 92 |
| Yes | 0 | 0 | 0 | 0 | 13 | 1 | 33 | 6 | 43 | 8 |
| By ATC-codes: C10AA, C10AB, C10AX (%) | | | | | | | | | | |
| No | 78 | 100 | 48 | 98 | 34 | 95 | 21 | 87 | 15 | 82 |
| Yes | 22 | 0 | 52 | 2 | 66 | 5 | 79 | 13 | 85 | 18 |
| **Diabetes** | **Yes** | **No** | **Yes** | **No** | **Yes** | **No** | **Yes** | **No** | **Yes** | **No** |
| Participants (N) | 180 | 5715 | 300 | 4055 | 446 | 3655 | 593 | 2989 | 639 | 2534 |
| By ICD-codes: E10, E11, E13, E14 (%) | | | | | | | | | | |
| No | 81 | 100 | 77 | 100 | 25 | 99 | 10 | 99 | 6 | 99 |
| Yes | 19 | 0 | 23 | 0 | 75 | 1 | 90 | 1 | 94 | 1 |
| By ATC-codes: A10A, A10B (%) | | | | | | | | | | |

| | Phase I | | Phase II | | Phase III | | Phase IV | | Phase V | |
|---|---|---|---|---|---|---|---|---|---|---|
| No | 36 | 100 | 20 | 99 | 6 | 99 | 4 | 99 | 3 | 98 |
| Yes | 64 | 0 | 80 | 1 | 94 | 1 | 96 | 1 | 97 | 2 |
| **Cardiovascular outcomes** | **Phase I** | | **Phase II** | | **Phase III** | | **Phase IV** | | **Phase V** | |
| **Heart attack** | **Yes** | **No** | **Yes** | **No** | **Yes** | **No** | **Yes** | **No** | **Yes** | **No** |
| Participants (N) | 39 | 5726 | 68 | 4236 | 82 | 3934 | NA | NA | NA | NA |
| By ICD-codes: I21 (%) | | | | | | | | | | |
| No | 87 | 100 | 68 | 100 | 48 | 100 | NA | NA | NA | NA |
| Yes | 13 | 0 | 32 | 0 | 52 | 0 | NA | NA | NA | NA |
| **Cerebrovascular disease** | **Yes** | **No** | **Yes** | **No** | **Yes** | **No** | **Yes** | **No** | **Yes** | **No** |
| Participants (N) | 89 | 5683 | - | 4203 | - | 3873 | NA | NA | 202 | 2828 |
| By ICD-codes: I60–I69 (%) | | | | | | | | | | |
| No | 20 | 38 | 3 | 16 | 1 | 6 | NA | NA | 0 | 0 |
| Yes | 80 | 61 | 97 | 83 | 99 | 94 | NA | NA | 100 | 100 |
| **Angina pectoris** | **Yes** | **No** | **Yes** | **No** | **Yes** | **No** | **Yes** | **No** | **Yes** | **No** |
| Participants (N) | 349 | 5436 | 115 | 4197 | 111 | 3903 | NA | NA | NA | NA |
| By ICD-codes: I20 (%) | | | | | | | | | | |
| No | 91 | 100 | 63 | 99 | 59 | 98 | NA | NA | NA | NA |
| Yes | 9 | 0 | 37 | 1 | 41 | 2 | NA | NA | NA | NA |
| **Intermittent claudication** | **Yes** | **No** | **Yes** | **No** | **Yes** | **No** | **Yes** | **No** | **Yes** | **No** |
| Participants (N) | 14 | 5744 | 22 | 4262 | - | 3968 | NA | NA | NA | NA |
| By ICD-codes: I73.9 (%) | | | | | | | | | | |
| No | 100 | 100 | 100 | 100 | 95 | 100 | NA | NA | NA | NA |
| Yes | 0 | 0 | 0 | 0 | 5 | 0 | NA | NA | NA | NA |

ICD: International Classification of Diseases (ICD-10)

ATC: Anatomical Therapeutic Chemical (ATC) codes

### Validation Table 3: Prevalence ratio and AUC between register-based and survey-based diagnoses

Validation Table 3. The prevalence of register-based measurements compared to survey-based measurements among the participants of the Helsinki Health Study who consented for register linkage. Age-adjusted binomial regression analysis (prevalence ratios [PR] and 95% confidence intervals [CI]). AUC = area under the (receiver operating characteristic [ROC]) curve. ROC curve = receiver operating characteristic curve.

| **Phases** | **Pooled Phase I-V** | | | |
|---|---|---|---|---|
| **<u>Metabolic risk factors</u>** | **PR** | **95% CI** | **AUC** | **ROC curve** |
| **Hypertension** | | | | |
| By ICD-codes | 21.31 | 18.77-24.18 | 0.94 | 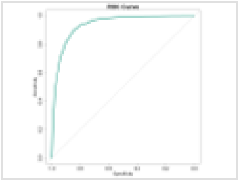 |
| By ATC-codes | 5.79 | 5.57-6.01 | 0.92 | 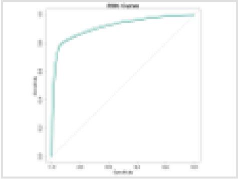 |
| **Hypercholesterolemia** | | | | |
| By ICD-codes | 5.82 | 5.16-6.57 | 0.90 | 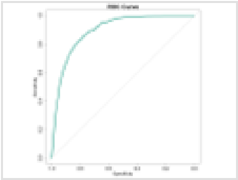 |
| By ATC-codes | 10.13 | 9.44-10.86 | 0.92 | 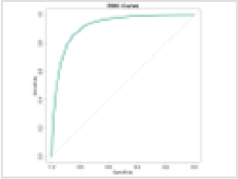 |
| **Diabetes** | | | | |

| | | | | |
|---|---|---|---|---|
| By ICD-codes | 123.33 | 100.69-151.06 | 0.98 | 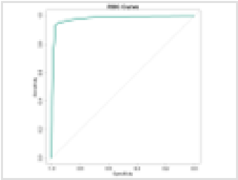 |
| By ATC-codes | 87.22 | 75.58-100.65 | 0.97 | 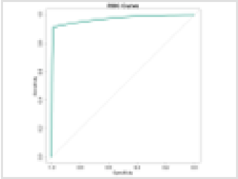 |
| **<u>Cardiovascular outcomes</u>** | **PR** | **95% CI** | **AUC** | |
| **Heart attack** | | | | |
| By ICD-codes | 4317.64 | 597.41-31204.43 | 0.99 | 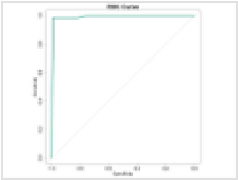 |
| **Cerebrovascular disorder** | | | | |
| By ICD-codes | 1.00 | 0.99-1.01 | 0.71 | 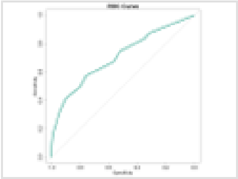 |
| **Angina pectoris** | | | | |
| By ICD-codes | 19.45 | 15.44-24.52 | 0.87 | 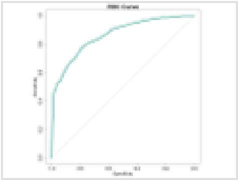 |
| **Claudication** | | | | |

| | | | | |
|---|---|---|---|---|
| By ICD-codes | 988495950.76 | 0.00-Inf | 1.00 | 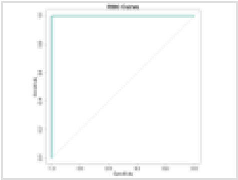 |

#### Validation Table 4: Reliability of survey-based diagnoses by starting phase

Validation Table 4. Reliability (percentage range) following initial positive 'yes' report in later phases with continued positive 'yes' responses. NA = item not measured in subsequent surveys.

| **Initial phase of measuring = Yes** | **Phase 1 (%-range)** | **Phase 2 (%-range)** | **Phase 3 (%-range)** | **Phase 4 (%-range)** |
|---|---|---|---|---|
| **<u>Metabolic risk factors</u>** | | | | |
| Hypertension | 89-95 | 92-96 | 95-96 | 97 |
| Hypercholesterolemia | 83-86 | 80-86 | 80 | NA |
| Diabetes | 92-93 | 95 | 96 | NA |
| **<u>Cardiovascular outcome</u>** | | | | |
| Angina pectoris | 17-19 | 52 | NA | NA |
| Heart attack | 68 | 87 | NA | NA |
| Claudication | 22-70 | 61 | NA | NA |
| Cerebrovascular disease | 49-67 | 71-78 | 71 | NA |

### 2. Other supplementary tables

##### Supplementary Table S1: Expanded Table 1 with summaries by each social isolation exposure separately

Supplementary Table S1. Summaries of the studied variables by all social isolation exposures and study inclusion. Data consisted of 40–60-year-old employees (n = 8 967) of the City of Helsinki, Finland, who responded to a Phase 1 survey in 2000-2002 (1) and 0-4 follow-up surveys in 2007 (2), 2012, 2017, or 2022 (3-5), out of which 5 403 were included to the main analysis.

| Variable | Summary | | Living alone | | Meeting friends/relatives | | Marital status | | Social support | | |
|---|---|---|---|---|---|---|---|---|---|---|---|
| | | Included | Yes | No | Rare | Often | Singles | Couples | 0-4 points | 5+ points | Excluded |
| **Total** | | | | | | | | | | | |
| | N | 5403 | 2182 | 6717 | 1431 | 6952 | 2728 | 6176 | 3471 | 5413 | 3564 |
| **Background** | | | | | | | | | | | |
| Age | (1) Years, mean | 48.8 | 51.5 | 48.7 | 49.3 | 49.4 | 49.5 | 49.3 | 50.4 | 48.7 | 50.2 |
| Occupational class | (1) Manual, # (%) | 1737 (32.1) | 714 (33.6) | 2235 (33.9) | 454 (32.4) | 2361 (34.6) | 1017 (38.2) | 1938 (31.9) | 1193 (35.3) | 1755 (32.9) | 1236 (36.6) |
| | (1) Routine non-manual, # (%) | 1164 (21.5) | 413 (19.4) | 1296 (19.6) | 246 (17.6) | 1368 (20.0) | 475 (17.9) | 1233 (20.3) | 579 (17.1) | 1125 (21.1) | 551 (16.3) |
| | (1) Semi-professional, # (%) | 1767 (32.7) | 584 (27.4) | 2010 (30.5) | 445 (31.8) | 2001 (29.3) | 654 (24.6) | 1935 (31.9) | 816 (24.1) | 1776 (33.3) | 833 (24.7) |
| | (1) Manager/professional, # (%) | 735 (13.6) | 417 (19.6) | 1059 (16.0) | 255 (18.2) | 1102 (16.1) | 515 (19.4) | 966 (15.9) | 794 (23.5) | 674 (12.6) | 757 (22.4) |
| Sex | Female, # (%) | 4365 (80.8) | 1726 (79.1) | 5394 (80.3) | 1102 (77.0) | 5594 (80.5) | 2332 (85.5) | 4796 (77.7) | 2623 (75.6) | 4491 (83.0) | 2803 (78.8) |
| **Exposures: Social isolation** | | | | | | | | | | | |
| Friends/Relatives | (1) < 2 meetups/month, # (%) | 850 (16.6) | 298 (14.7) | 1128 (17.9) | 1431 (100.0) | 0 (0.0) | 317 (12.5) | 1108 (19.1) | 718 (22.1) | 712 (13.9) | 581 (17.9) |
| Living alone | (1) Yes, # (%) | 1222 (22.7) | 2182 (100.0) | 0 (0.0) | 298 (20.9) | 1733 (25.1) | 1587 (59.0) | 578 (9.4) | 988 (28.7) | 1172 (21.8) | 960 (27.3) |
| Marital status | (1) Single, # (%) | 1569 (29.2) | 1587 (73.3) | 1105 (16.5) | 317 (22.2) | 2213 (32.0) | 2728 (100.0) | 0 (0.0) | 1257 (36.5) | 1443 (26.8) | 1159 (32.8) |
| Social support | (1) Score 0-4, # (%) | 1886 (35.1) | 988 (45.7) | 2454 (36.8) | 718 (50.2) | 2528 (36.4) | 1257 (46.6) | 2189 (35.7) | 3471 (100.0) | 0 (0.0) | 1585 (45.2) |
| **Mediators: Metabolic factors** | | | | | | | | | | | |
| Diabetes | (1) Has history, # (%) | 112 (2.2) | 71 (3.8) | 166 (2.8) | 43 (3.4) | 175 (2.8) | 88 (3.7) | 148 (2.7) | 111 (3.7) | 120 (2.5) | 125 (4.5) |
| | (2) New, # (%) | 143 (3.1) | 53 (4.6) | 149 (3.8) | 27 (3.4) | 165 (4.1) | 57 (3.9) | 145 (4.1) | 80 (4.5) | 121 (3.7) | 60 (14.3) |
| Hypercholesterolemia | (1) Has history, # (%) | 1039 (19.3) | 542 (25.1) | 1331 (20.0) | 301 (21.2) | 1464 (21.2) | 585 (21.6) | 1287 (21.0) | 761 (22.2) | 1111 (20.7) | 846 (24.2) |
| | (2) New, # (%) | 863 (20.6) | 306 (23.6) | 891 (20.7) | 210 (23.4) | 939 (21.3) | 363 (21.7) | 833 (21.2) | 465 (22.2) | 727 (20.8) | 340 (23.5) |
| Hypertension | (1) Has history, # (%) | 1067 (19.8) | 562 (26.0) | 1457 (21.9) | 340 (24.0) | 1545 (22.4) | 589 (21.8) | 1436 (23.5) | 839 (24.4) | 1178 (22.0) | 964 (27.7) |
| | (2) New, # (%) | 602 (14.6) | 219 (17.1) | 673 (16.0) | 149 (17.1) | 713 (16.4) | 262 (15.7) | 628 (16.5) | 379 (18.6) | 512 (14.9) | 299 (21.7) |
| Obesity (BMI 30+) | (1) Yes, # (%) | 677 (12.6) | 346 (16.2) | 920 (13.8) | 211 (15.0) | 990 (14.4) | 423 (15.7) | 844 (13.8) | 564 (16.5) | 703 (13.1) | 598 (17.2) |
| | (2) Changed, # (%) | 410 (7.9) | 177 (10.2) | 426 (7.9) | 102 (9.0) | 469 (8.3) | 191 (8.9) | 411 (8.2) | 249 (9.2) | 354 (8.0) | 195 (9.7) |
| **Mediators: Health behaviours** | | | | | | | | | | | |
| Alcohol consumption | (1) > 7/14 units/week, # (%) | 860 (16.3) | 361 (17.5) | 1043 (16.2) | 251 (18.2) | 1081 (16.2) | 413 (16.0) | 989 (16.7) | 497 (15.2) | 907 (17.3) | 551 (16.8) |
| | (2) Changed, # (%) | 628 (12.5) | 189 (11.6) | 665 (12.9) | 138 (12.6) | 667 (12.4) | 234 (11.8) | 620 (12.9) | 301 (11.9) | 554 (13.0) | 229 (12.9) |
| Fruit & vegetable consumption | (1) < 1 serving/day, # (%) | 641 (11.9) | 354 (16.4) | 822 (12.3) | 242 (17.0) | 868 (12.6) | 473 (17.6) | 696 (11.4) | 563 (16.4) | 607 (11.3) | 540 (15.5) |
| | (2) Changed, # (%) | 603 (11.7) | 235 (13.6) | 644 (11.9) | 165 (14.5) | 662 (11.8) | 317 (15.0) | 561 (11.2) | 387 (14.4) | 489 (11.0) | 283 (14.3) |
| Physical activity | (1) < 14 MET-hours/week, # (%) | 1235 (23.0) | 545 (25.4) | 1678 (25.2) | 435 (30.6) | 1661 (24.1) | 712 (26.5) | 1513 (24.7) | 1016 (29.6) | 1209 (22.5) | 1002 (28.6) |
| | (2) Changed, # (%) | 1212 (23.4) | 429 (24.8) | 1341 (24.7) | 313 (27.1) | 1373 (24.3) | 545 (25.5) | 1227 (24.5) | 742 (27.5) | 1030 (23.1) | 568 (28.2) |
| Sleep duration | (1) < 7 hours/workday, # (%) | 1196 (22.4) | 562 (26.2) | 1528 (23.2) | 374 (26.7) | 1578 (23.1) | 701 (26.1) | 1382 (22.8) | 911 (26.7) | 1174 (22.1) | 906 (26.3) |
| | (2) Changed, # (%) | 1029 (20.0) | 358 (20.8) | 1097 (20.5) | 241 (21.3) | 1135 (20.4) | 436 (20.5) | 1008 (20.4) | 585 (21.7) | 867 (19.8) | 431 (21.9) |
| Smoking | (1) Current regular, # (%) | 1123 (20.9) | 547 (25.3) | 1552 (23.3) | 315 (22.2) | 1652 (24.0) | 792 (29.5) | 1308 (21.3) | 893 (26.1) | 1208 (22.5) | 993 (28.4) |
| | (2) Changed, # (%) | 389 (7.5) | 107 (6.1) | 430 (7.9) | 75 (6.5) | 432 (7.6) | 167 (7.8) | 369 (7.4) | 204 (7.6) | 331 (7.4) | 151 (7.5) |
| **Outcomes: CVD** | | | | | | | | | | | |
| Cardiovascular disease | (3-5) Yes, # (%) | 993 (18.4) | 481 (22.0) | 1267 (18.9) | 292 (20.4) | 1365 (19.6) | 550 (20.2) | 1200 (19.4) | 705 (20.3) | 1037 (19.2) | 765 (21.5) |
| Lost to follow-up | (3-5) Yes, # (%) | 2133 (39.5) | 1120 (51.3) | 3183 (47.4) | 684 (47.8) | 3307 (47.6) | 1387 (50.8) | 2914 (47.2) | 1797 (51.8) | 2493 (46.1) | 2210 (62.0) |
| Rose questionnaire only | (3-5) Yes, # (%) | 356 (6.6) | 173 (7.9) | 422 (6.3) | 98 (6.8) | 470 (6.8) | 232 (8.5) | 362 (5.9) | 233 (6.7) | 359 (6.6) | 240 (6.7) |

### Supplementary table S2: Direct and indirect effects in mediation analysis

| Mediator | Pathway | Exposure | | | | |
|---|---|---|---|---|---|---|
| | | Social isolation (all vs. none) | Living alone | Rare meetups (< 2/month) | Low support (0-4 points) | Single (marital status) |
| **Metabolic factors** | | | | | | |
| Hypertension | Direct | 1.00 (0.75-1.27) | 1.10 (0.92-1.30) | 0.93 (0.76-1.11) | 0.96 (0.85-1.09) | 1.01 (0.85-1.20) |
| | Indirect | 1.00 (0.99-1.02) | 1.00 (0.99-1.01) | 1.00 (0.99-1.01) | 1.00 (0.99-1.01) | 1.00 (0.99-1.01) |
| Diabetes | Direct | 1.00 (0.75-1.28) | 1.11 (0.93-1.32) | 0.94 (0.77-1.13) | 0.96 (0.85-1.09) | 0.99 (0.83-1.17) |
| | Indirect | 1.00 (0.99-1.01) | 1.01 (1.00-1.02) | 1.00 (0.99-1.00) | 1.00 (0.99-1.00) | 1.00 (0.99-1.01) |
| Hypercholesterolemia | Direct | 1.00 (0.76-1.28) | 1.10 (0.92-1.30) | 0.94 (0.77-1.13) | 0.97 (0.85-1.10) | 1.00 (0.84-1.18) |
| | Indirect | 1.00 (1.00-1.01) | 1.00 (1.00-1.01) | 1.00 (1.00-1.01) | 1.00 (0.99-1.00) | 1.00 (0.99-1.00) |
| Obesity (BMI 30+) | Direct | 1.00 (0.76-1.27) | 1.12 (0.93-1.32) | 0.94 (0.77-1.13) | 0.96 (0.84-1.09) | 0.99 (0.83-1.17) |
| | Indirect | 1.00 (0.99-1.01) | 1.01 (1.00-1.01) | 1.00 (0.99-1.00) | 1.00 (1.00-1.01) | 1.00 (0.99-1.00) |
| **Health behaviors** | | | | | | |
| Smoking | Direct | 1.00 (0.77-1.28) | 1.13 (0.94-1.33) | 0.96 (0.78-1.14) | 0.96 (0.84-1.09) | 0.98 (0.82-1.15) |
| | Indirect | 1.00 (0.99-1.01) | 1.00 (1.00-1.00) | 1.00 (1.00-1.00) | 1.00 (1.00-1.00) | 1.00 (0.99-1.00) |
| Heavy alcohol consumption | Direct | 1.01 (0.76-1.29) | 1.11 (0.93-1.32) | 0.95 (0.78-1.14) | 0.96 (0.84-1.09) | 1.00 (0.84-1.18) |
| | Indirect | 1.00 (1.00-1.01) | 1.00 (1.00-1.01) | 1.00 (0.99-1.00) | 1.00 (1.00-1.00) | 1.00 (1.00-1.00) |
| Short sleep duration | Direct | 0.98 (0.74-1.25) | 1.11 (0.93-1.32) | 0.94 (0.76-1.12) | 0.95 (0.84-1.08) | 0.99 (0.83-1.18) |
| | Indirect | 1.00 (1.00-1.01) | 1.00 (1.00-1.00) | 1.00 (1.00-1.00) | 1.00 (1.00-1.00) | 1.00 (1.00-1.01) |
| Low fruit & vegetable consumption | Direct | 1.00 (0.76-1.27) | 1.12 (0.94-1.33) | 0.94 (0.77-1.12) | 0.96 (0.85-1.08) | 0.99 (0.82-1.16) |
| | Indirect | 1.01 (1.00-1.03) | 1.00 (0.99-1.00) | 1.00 (1.00-1.01) | 1.00 (1.00-1.01) | 1.01 (1.00-1.01) |
| Physical inactivity | Direct | 0.93 (0.71-1.18) | 1.11 (0.93-1.32) | 0.91 (0.74-1.08) | 0.94 (0.83-1.06) | 0.99 (0.83-1.17) |
| | Indirect | 1.02 (1.00-1.04) | 1.00 (1.00-1.01) | 1.01 (1.00-1.01) | 1.00 (1.00-1.01) | 1.00 (1.00-1.01) |
| **Mediator composites** | | | | | | |
| 2+ metabolic factors | Direct | 0.99 (0.75-1.27) | 1.10 (0.92-1.30) | 0.94 (0.77-1.12) | 0.97 (0.85-1.09) | 1.00 (0.84-1.18) |
| | Indirect | 1.00 (0.99-1.02) | 1.01 (1.00-1.02) | 1.00 (0.99-1.01) | 1.00 (0.99-1.01) | 0.99 (0.98-1.01) |
| 2+ behavioral factors | Direct | 0.96 (0.73-1.22) | 1.12 (0.93-1.32) | 0.92 (0.76-1.10) | 0.95 (0.83-1.08) | 0.98 (0.82-1.16) |
| | Indirect | 1.00 (0.99-1.01) | 1.00 (1.00-1.00) | 1.00 (1.00-1.01) | 1.00 (1.00-1.00) | 1.00 (1.00-1.01) |
| 4+ risk factors | Direct | 0.95 (0.72-1.22) | 1.11 (0.92-1.31) | 0.92 (0.75-1.11) | 0.95 (0.83-1.08) | 0.99 (0.82-1.17) |
| | Indirect | 1.01 (1.00-1.02) | 1.00 (1.00-1.01) | 1.00 (1.00-1.01) | 1.00 (1.00-1.00) | 1.00 (1.00-1.01) |
| **Multi-mediators models** | | | | | | |
| Behavioral factors | Direct | 0.92 (0.70-1.17) | 1.11 (0.93-1.32) | 0.91 (0.75-1.09) | 0.94 (0.82-1.06) | 0.97 (0.81-1.15) |
| | Indirect | 1.03 (1.01-1.05) | 1.01 (1.00-1.02) | 1.01 (1.00-1.02) | 1.01 (1.00-1.02) | 1.01 (1.00-1.02) |
| Metabolic factors | Direct | 0.98 (0.75-1.25) | 1.08 (0.91-1.28) | 0.93 (0.76-1.11) | 0.97 (0.85-1.09) | 1.01 (0.85-1.20) |
| | Indirect | 1.00 (0.99-1.02) | 1.01 (1.00-1.03) | 1.00 (0.99-1.01) | 1.00 (0.99-1.01) | 0.99 (0.98-1.00) |
| All factors | Direct | 0.91 (0.70-1.16) | 1.09 (0.91-1.28) | 0.91 (0.75-1.08) | 0.95 (0.84-1.07) | 0.99 (0.83-1.17) |
| | Indirect | 1.03 (1.00-1.05) | 1.02 (1.00-1.03) | 1.01 (0.99-1.02) | 1.01 (0.99-1.02) | 1.00 (0.98-1.01) |

Supplementary Table S2. Estimated direct and indirect effects (risk ratios and 90% credible intervals) of social isolation and each social isolation exposure on the risk of cardiovascular disease through each mediator separately, through mediator composite variables, and multiple mediators modelled jointly. All analyses were adjusted for age, sex, occupational class, as well as for the Phase 1 value of the mediators. The data consisted of 10,175 observations from 5,403 participants in the Helsinki Health Study of employees of the City of Helsinki, Finland.

### 3. Supplementary figures

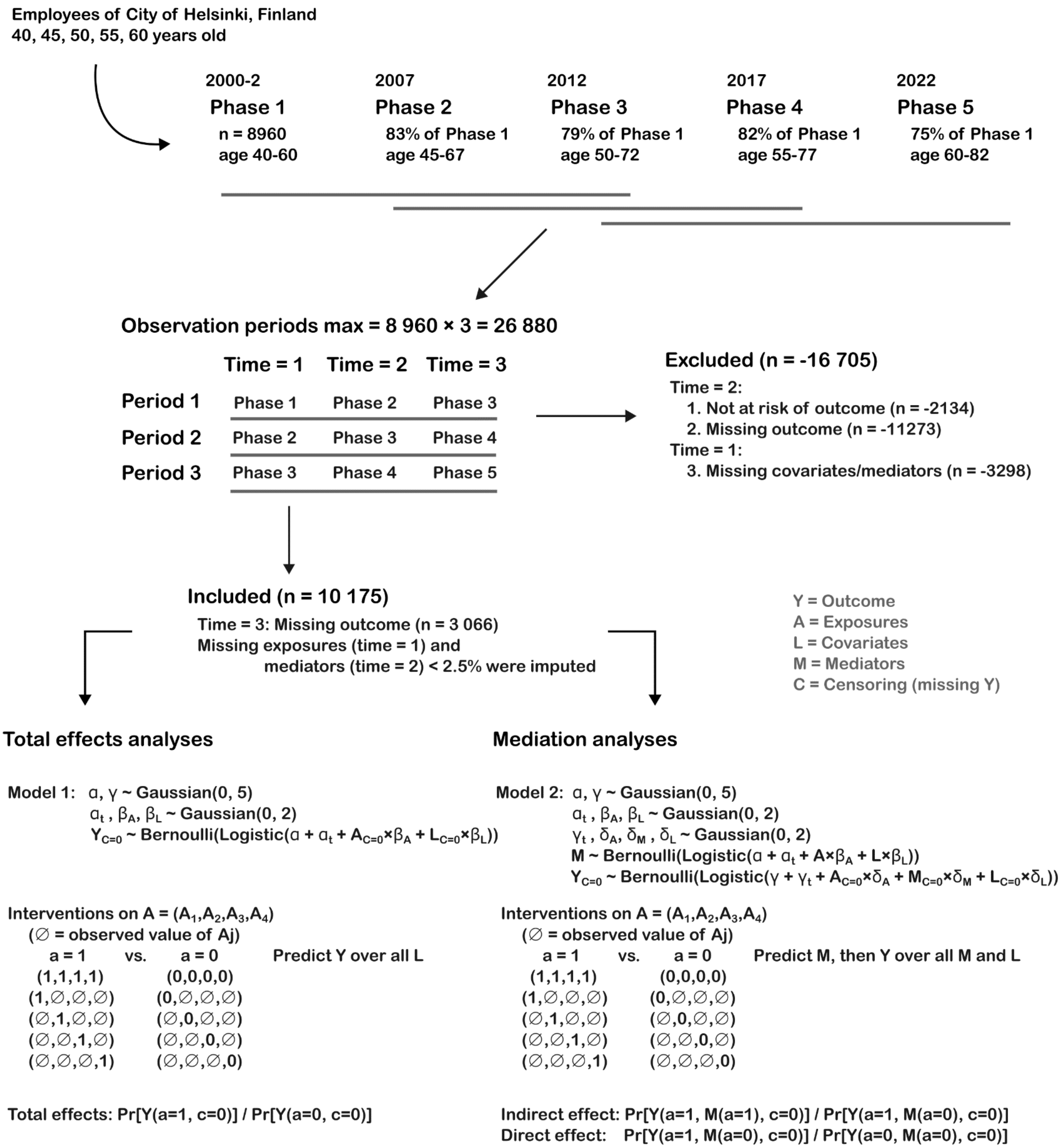

Supplementary Figure S1. A visual illustration of the flow of the study from the Phase 1 survey in 2000-2 to the estimates of total, indirect, and direct effects.

### 4. R and Stan code

```r
# Tip: Search for "run_analysis" for the main function and
# "stanmodel <-" for the Stan models.

library(tidyverse)
library(rstan)
library(flextable)

options(mc.cores = parallel::detectCores())

import_sas <- function(datadir, varname_map) {
    filenames <- dir(path = datadir)
    fullpaths <- list.files(path = datadir, full.names = TRUE)
    waves <- parse_number(filenames) # First number in file name is phase.

    dflist <- map2(
        fullpaths, waves,
        \(p,w) haven::read_sas(p) |>
            # Remove SAS-metadata.
            haven::zap_label() |>
            haven::zap_formats() |>
            haven::zap_labels() |>
            haven::zap_missing() |>
            haven::zap_widths() |>
            mutate(across(where(is.character), haven::zap_empty)) |>
            # All variables are coded numerically.
            mutate(across(everything(), as.numeric)) |>
            rename(any_of(varname_map)) |>
            mutate(id = as.character(id), wave = w)
    )

    df <- dflist |>
        bind_rows() |>
        arrange(id, wave) |>
        complete(id, wave) |>
        select(id, wave, sort(tidyselect::peek_vars()))

    return(df)
}

transform_sex <- function(df) {
    df |>
        # Fill sex to other time points from the 1st survey.
        # Needed for defining heavy alcohol consumption.
        group_by(id) |>
        mutate(sex = if_else(sex == 2, true = 0, false = 1)) |>
        fill(sex, .direction = "down") |>
        ungroup()
}

transform_alco <- function(df) {
    df |>
        mutate(
            # From categorical answers to average units.
            alcoa = case_when(
```

```r
                alcoa == 1 ~ 0,
                alcoa == 2 ~ 0.5,
                alcoa == 3 ~ 2.5,
                alcoa == 4 ~ 8.5,
                alcoa == 5 ~ 18.5,
                alcoa == 6 ~ 36,
                alcoa == 7 ~ 48,
                TRUE ~ NA_real_
            ),
            alcob = case_when(
                alcob == 1 ~ 0,
                alcob == 2 ~ 0.5,
                alcob == 3 ~ 2.5,
                alcob == 4 ~ 10.5,
                alcob == 5 ~ 22.5,
                alcob == 6 ~ 42,
                alcob == 7 ~ 60,
                TRUE ~ NA_real_
            ),
            alcoc = case_when(
                alcoc == 1 ~ 0,
                alcoc == 2 ~ 0.5,
                alcoc == 3 ~ 3,
                alcoc == 4 ~ 9,
                alcoc == 5 ~ 20,
                alcoc == 6 ~ 47,
                alcoc == 7 ~ 65,
                TRUE ~ NA_real_
            ),
            alco = alcoa + alcob + alcoc,
            # From units to moderate vs. heavy consumption.
            alco01 = case_when(
                sex == 0 & alco >= 7 ~ 1L,
                sex == 0 & alco < 7 ~ 0L,
                sex == 1 & alco >= 14 ~ 1L,
                sex == 1 & alco < 14 ~ 0L,
                alco < 7 ~ 0L,
                TRUE ~ NA_integer_
            )
        ) |>
        select(-alcoa, -alcob, -alcoc, -alcohabit)
}

transform_smoking <- function(df) {
    df |>
        mutate(smoker = if_else(smoker == 2, true = 0, false = 1)) |>
        # `rowwise` is needed because `sum` is not vectorized as needed.
        rowwise() |>
        mutate(
            smoking = case_when(
                is.na(smoking123) &&
                    any(!is.na(c(smoking1, smoking2, smoking3))) ~
                        sum(c(smoking1, smoking2, smoking3), na.rm = TRUE),
                is.na(smoking123) & smoker == 0 ~ 0,
                TRUE ~ smoking123
            )
```

```r
        ) |>
        ungroup() |>
        select(-matches("smoking[0-9]")) |>
        mutate(
            smoker = if_else(
                condition = is.na(smoker) & smoking > 0,
                true = 1,
                false = smoker
            )
        )
}

transform_cohab <- function(df) {
    df |>
        # Note: Counts are truncated to 9 or 5 depending on survey.
        mutate(
            cohab2 = if_else(cohab2 == 2, true = 0, false = 1)
            # Other cohabitation items are already counts from 0.
        ) |>
        rowwise() |>
        mutate(
            # Add children together when reported separately.
            cohab34 = case_when(
                is.na(cohab34) && any(!is.na(c(cohab3, cohab4))) ~
                    sum(c(cohab3, cohab4), na.rm = TRUE),
                TRUE ~ cohab34
            ),
            # Add all counts together ignoring whether child/adult.
            cohabs = case_when(
                cohab1 == 2 ~ 0L,
                # If any sub-answer given, others assumed 0.
                any(!is.na(c(cohab2, cohab34, cohab5))) ~
                    sum(c(cohab2, cohab34, cohab5), na.rm = TRUE),
                TRUE ~ NA_integer_
            ),
            livingalone = if_else(cohabs == 0, 1L, 0L)
        ) |>
        ungroup()
}

transform_support <- function(df) {
    df |>
        # Note: In surveys 1-4, all-zeros means item-nonresponse
        # and NA is survey-nonresponse, but in the 5th survey, ticks are 1
        # and non-ticks are missing.
        rowwise() |>
        mutate(
            support_all_a = case_when(
                # Case 1: Item-nonresponse means missing.
                all(c_across(matches("support_a[1-7]")) == 0) ~ NA_integer_,
                # Case 2: 'No-one' means the sum is 0.
                support_a7 == 1 ~ 0,
                # Case 3: 5th survey can have ones and NAs; then NA means '0'.
                all(c(NA, 1) %in% unique(c_across(matches("support_a[1-7]")))) ~
                    sum(c_across(matches("support_a[1-6]")), na.rm = TRUE),
                # Case 4: Otherwise sum points with na.rm = FALSE.
```

```
                TRUE ~ sum(c_across(matches("support_a[1-6]")), na.rm = FALSE)
            ),
            support_all_b = case_when(
                all(c_across(matches("support_b[1-7]")) == 0) ~ NA_integer_,
                support_b7 == 1 ~ 0,
                all(c(NA, 1) %in% unique(c_across(matches("support_b[1-7]")))) ~
                    sum(c_across(matches("support_b[1-6]")), na.rm = TRUE),
                TRUE ~ sum(c_across(matches("support_b[1-6]")), na.rm = FALSE)
            ),
            support_all_c = case_when(
                all(c_across(matches("support_c[1-7]")) == 0) ~ NA_integer_,
                support_c7 == 1 ~ 0,
                all(c(NA, 1) %in% unique(c_across(matches("support_c[1-7]")))) ~
                    sum(c_across(matches("support_c[1-6]")), na.rm = TRUE),
                TRUE ~ sum(c_across(matches("support_c[1-6]")), na.rm = FALSE)
            ),
            support_all_d = case_when(
                all(c_across(matches("support_d[1-7]")) == 0) ~ NA_integer_,
                support_d7 == 1 ~ 0,
                all(c(NA, 1) %in% unique(c_across(matches("support_d[1-7]")))) ~
                    sum(c_across(matches("support_d[1-6]")), na.rm = TRUE),
                TRUE ~ sum(c_across(matches("support_d[1-6]")), na.rm = FALSE)
            ),
            # If any answers given in a-d, assume 'no-one' (0) for the missing a-d.
            support_all = case_when(
                any(!is.na(c_across(starts_with("support_all")))) ~
                    sum(c_across(starts_with("support_all")), na.rm = TRUE),
                TRUE ~ NA_integer_
            )
        ) |>
        ungroup() |>
        # Low social support defined as 0-4 points.
        mutate(support01 = if_else(support_all <= 4, 1L, 0L)) |>
        select(-matches("support_[a-d][1-7]"))
}

transform_meetups <- function(df) {
    df |>
        mutate(
            relatmeets_count = case_when(
                relatmeets == 1 ~ 20,
                relatmeets == 2 ~ 4,
                relatmeets == 3 ~ 1,
                relatmeets == 4 ~ 1/4.5,
                relatmeets %in% 5:6 ~ 0,
                TRUE ~ NA_real_
            ),
            friendmeets_count = case_when(
                friendmeets == 1 ~ 20,
                friendmeets == 2 ~ 4,
                friendmeets == 3 ~ 1,
                friendmeets == 4 ~ 1/4.5,
                friendmeets == 5 ~ 0,
                TRUE ~ NA_real_
            ),
            totalmeets = relatmeets_count + friendmeets_count,
```

```
            raremeets = if_else(totalmeets < 2, true = 1L, false = 0L)
        )
}

transform_fruveg <- function(df) {
    df |>
        mutate(
            # From categories to average number of servings.
            fruit4wk = case_when(
                fruit == 1 ~ 0,
                fruit == 2 ~ 2,
                fruit == 3 ~ 4,
                fruit == 4 ~ 12,
                fruit == 5 ~ 22,
                fruit == 6 ~ 28,
                fruit == 7 ~ 56,
                TRUE ~ NA_integer_
            ),
            veg4wk = case_when(
                veget == 1 ~ 0,
                veget == 2 ~ 2,
                veget == 3 ~ 4,
                veget == 4 ~ 12,
                veget == 5 ~ 22,
                veget == 6 ~ 28,
                veget == 7 ~ 56,
                TRUE ~ NA_integer_
            ),
            fruveg = fruit4wk + veg4wk,
            # Sub-daily fruit or vegetables.
            fruveg01 = if_else(fruveg < 28, true = 1L, false = 0L)
        ) |>
        select(-fruit4wk, -veg4wk, -veget, -fruit)
}

tidy_sas <- function(df) {
    df |>
        transform_sex() |>
        # Social isolation.
        mutate(
            # Note "1 = more isolation" coding throughout.
            civstat01 = case_when(
                # Registered partnership, married.
                civstat %in% c(2, 3) ~ 0L,
                # Unmarried, separated/divorced, widow.
                civstat %in% c(1, 4, 5) ~ 1L,
                TRUE ~ NA_integer_
            )
        ) |>
        transform_support() |>
        transform_cohab() |>
        transform_meetups() |>
        # Health behaviours.
        transform_alco() |>
        transform_smoking() |>
        transform_fruveg() |>
```

```
    mutate(
        sleephr = sleephr + 4,
        sleep01 = if_else(sleephr >= 7, true = 0L, false = 1L),
        met01 = if_else(met < 14, true = 1L, false = 0L)
    ) |>
    # Metabolic syndrome.
    mutate(
        hbp = if_else(hbp == 2, true = 0, false = 1),
        hc = if_else(hc == 2, true = 0, false = 1),
        diab = if_else(diab == 2, true = 0, false = 1),
        bmi01 = if_else(bmi >= 30, true = 1, false = 0),
        metabany01 = case_when(
            hbp == 1 | hc == 1 | diab == 1 | bmi01 == 1 ~ 1L,
            hbp == 0 & hc == 0 & diab == 0 & bmi01 == 0 ~ 0L,
            TRUE ~ NA_integer_
        )
    ) |>
    # Mediator composites.
    rowwise() |>
    mutate(
        # Some missing values can be allowed as long as ">= 2" is certain.
        behavmulti01 = case_when(
            # 2+ adverse behaviors observed: is "yes".
            c(met01, fruveg01, sleep01, smoker, alco01) |>
                sum(na.rm = TRUE) >= 2 ~ 1L,
            # 4+ non-adverse behaviors observed: cannot be "yes".
            (c(met01, fruveg01, sleep01, smoker, alco01) == 0) |>
                sum(na.rm = TRUE) >= 4 ~ 0L,
            # Otherwise it's unknown.
            TRUE ~ NA_integer_
        ),
        metabmulti01 = case_when(
            c(diab, hbp, hc, bmi01) |>
                sum(na.rm = TRUE) >= 2 ~ 1L,
            (c(diab, hbp, hc, bmi01) == 0) |>
                sum(na.rm = TRUE) >= 3 ~ 0L,
            TRUE ~ NA_integer_
        ),
        polyrisk01 = case_when(
            c(met01, fruveg01, sleep01, smoker, alco01, diab, hbp, hc, bmi01) |>
                sum(na.rm = TRUE) >= 4 ~ 1L,
            (c(met01, fruveg01, sleep01, smoker, alco01, diab, hbp, hc, bmi01) == 0) |>
                sum(na.rm = TRUE) >= 6 ~ 0L,
            TRUE ~ NA_integer_
        )
    ) |>
    ungroup() |>
    # CVD variables from (1=yes,2=no) to (0=no,1=yes).
    mutate(across(
        c(ami, cevd, angpect, chd, other_cvd, chd_other),
        \(x) if_else(x == 2, true = 0, false = x)
    )) |>
    # chd and other_cvd are measured in 4th survey.
    # chd_other in 5th survey combines them into one.
    # So we can add 4th survey information to chd_other.
    mutate(chd_other = case_when(
```

```r
            is.na(chd_other) & (chd == 0 & other_cvd == 0) ~ 0L,
            is.na(chd_other) & (chd == 1 | other_cvd == 1) ~ 1L,
            !is.na(chd_other) ~ chd_other,
            TRUE ~ NA_integer_
        ))
}

import_tidydata <- function(update,
                            tidyfile,
                            rawdir = NULL,
                            varnamefile = NULL) {
    if (update) {
        varname_map <- varnamefile |>
            read_csv(col_types = "cc") |>
            deframe()
        hhs <<- rawdir |>
            import_sas(varname_map) |>
            tidy_sas()
        save(hhs, file = tidyfile)
    } else {
        load(tidyfile, envir = globalenv())
    }
}

impute_logreg <- function(df, Y, X) {
    df_all <- df |> mutate(index = row_number())
    df_nona <- df_all |> drop_na()
    # Nothing to impute?
    if (nrow(df_all) == nrow(df_nona)) return(df)
    indeces_with_na <- which(is.na(df[[Y]]))
    df_yesna <- df_all |> filter(index %in% indeces_with_na)
    # Logistic regression model formula.
    frml <- as.formula(paste0(Y,"~1+",paste0(X,collapse="+")))
    model <- glm(formula = frml, data = df_nona, family = binomial(link = "logit"))
    # Note this is single-imputation but from a probability distribution.
    preds <- rbinom(
        n = length(indeces_with_na),
        size = 1,
        prob = predict.glm(model, newdata = df_yesna[ , X], type = "response")
    )
    df[indeces_with_na, Y] <- preds
    return(df)
}

# Function for dividing the five-phase person-time dataset into
# three observation periods with three time points.
prepare_dataset <- function(hhs) {

    df <- hhs |>
        select(
            id, wave,
            sex, age, sep,
            # Exposures.
            livingalone, raremeets, support01, civstat01,
            # Mediators.
            diab, hbp, hc, bmi01,
```

```r
        smoker, alco01, met01, fruveg01, sleep01,
        behavmulti01, metabmulti01, polyrisk01,
        # CVD outcome items (incl. inclusion criteria).
        ami, angpect, chd, rose, chd_other
    ) |>
    mutate(
        sep = if_else(sep == 11, 1L, as.integer(sep)),
        chdout = case_when(
            wave %in% 1:3 &
                (ami == 1 | angpect == 1 | rose == 1) ~ 1L,
            wave %in% 1:3 &
                (ami == 0 & angpect == 0 & rose == 0) ~ 0L,
            wave %in% 4:5 &
                (chd_other == 1 | rose == 1) ~ 1L,
            wave %in% 4:5 &
                (chd_other == 0 & rose == 0) ~ 0L,
            TRUE ~ NA_integer_
        )
    ) |>
    select(-(ami:chd_other)) |>
    # From person-time to wide-format.
    pivot_wider(names_from = wave, values_from = sex:chdout) |>
    # For ease of interpretation, you may pretend that observations
    # 1->2->3, 2->3->4, and 3->4->5 are three different (pseudo-)participants.
    # Below, "_1" or "_3" refer to these different pseudo-participants.
    select(
        id,

        sex_1, sex_2, sex_3,
        age_1, age_2, age_3,
        sep_1, sep_2, sep_3,

        livingalone_1, livingalone_2, livingalone_3,
        raremeets_1, raremeets_2, raremeets_3,
        support01_1, support01_2, support01_3,
        civstat01_1, civstat01_2, civstat01_3,

        # "bl" refers to baseline.
        chdout_bl_1 = chdout_2,
        chdout_bl_2 = chdout_3,
        chdout_bl_3 = chdout_4,
        chdout_1 = chdout_3,
        chdout_2 = chdout_4,
        chdout_3 = chdout_5,

        diab_bl_1 = diab_1,
        diab_bl_2 = diab_2,
        diab_bl_3 = diab_3,
        diab_1 = diab_2,
        diab_2 = diab_3,
        diab_3 = diab_4,
        hbp_bl_1 = hbp_1,
        hbp_bl_2 = hbp_2,
        hbp_bl_3 = hbp_3,
        hbp_1 = hbp_2,
        hbp_2 = hbp_3,
```

```
        hbp_3 = hbp_4,

        hc_bl_1 = hc_1,
        hc_bl_2 = hc_2,
        hc_bl_3 = hc_3,
        hc_1 = hc_2,
        hc_2 = hc_3,
        hc_3 = hc_4,

        bmi01_bl_1 = bmi01_1,
        bmi01_bl_2 = bmi01_2,
        bmi01_bl_3 = bmi01_3,
        bmi01_1 = bmi01_2,
        bmi01_2 = bmi01_3,
        bmi01_3 = bmi01_4,

        smoker_bl_1 = smoker_1,
        smoker_bl_2 = smoker_2,
        smoker_bl_3 = smoker_3,
        smoker_1 = smoker_2,
        smoker_2 = smoker_3,
        smoker_3 = smoker_4,

        alco01_bl_1 = alco01_1,
        alco01_bl_2 = alco01_2,
        alco01_bl_3 = alco01_3,
        alco01_1 = alco01_2,
        alco01_2 = alco01_3,
        alco01_3 = alco01_4,

        met01_bl_1 = met01_1,
        met01_bl_2 = met01_2,
        met01_bl_3 = met01_3,
        met01_1 = met01_2,
        met01_2 = met01_3,
        met01_3 = met01_4,

        fruveg01_bl_1 = fruveg01_1,
        fruveg01_bl_2 = fruveg01_2,
        fruveg01_bl_3 = fruveg01_3,
        fruveg01_1 = fruveg01_2,
        fruveg01_2 = fruveg01_3,
        fruveg01_3 = fruveg01_4,

        sleep01_bl_1 = sleep01_1,
        sleep01_bl_2 = sleep01_2,
        sleep01_bl_3 = sleep01_3,
        sleep01_1 = sleep01_2,
        sleep01_2 = sleep01_3,
        sleep01_3 = sleep01_4,

        behavmulti01_bl_1 = behavmulti01_1,
        behavmulti01_bl_2 = behavmulti01_2,
        behavmulti01_bl_3 = behavmulti01_3,
        behavmulti01_1 = behavmulti01_2,
        behavmulti01_2 = behavmulti01_3,
```

```
            behavmulti01_3 = behavmulti01_4,

            metabmulti01_bl_1 = metabmulti01_1,
            metabmulti01_bl_2 = metabmulti01_2,
            metabmulti01_bl_3 = metabmulti01_3,
            metabmulti01_1 = metabmulti01_2,
            metabmulti01_2 = metabmulti01_3,
            metabmulti01_3 = metabmulti01_4,

            polyrisk01_bl_1 = polyrisk01_1,
            polyrisk01_bl_2 = polyrisk01_2,
            polyrisk01_bl_3 = polyrisk01_3,
            polyrisk01_1 = polyrisk01_2,
            polyrisk01_2 = polyrisk01_3,
            polyrisk01_3 = polyrisk01_4
        ) |>
        mutate(
            # Impute known ages and socioeconomic positions.
            age_3 = case_when(
                !is.na(age_3) ~ age_3,
                is.na(age_3) & !is.na(age_2) ~ age_2 + 5,
                is.na(age_3) & is.na(age_2) & !is.na(age_1) ~ age_1 + 10,
                TRUE ~ NA_integer_
            ),
            age_2 = case_when(
                !is.na(age_2) ~ age_2,
                is.na(age_2) & !is.na(age_3) ~ age_3 - 5,
                is.na(age_2) & is.na(age_3) & !is.na(age_1) ~ age_1 + 5,
                TRUE ~ NA_integer_
            ),
            age_1 = case_when(
                !is.na(age_1) ~ age_1,
                is.na(age_1) & !is.na(age_2) ~ age_2 - 5,
                is.na(age_1) & is.na(age_2) & !is.na(age_3) ~ age_3 - 10,
                TRUE ~ NA_integer_
            ),
            sep_2 = sep_1,
            sep_3 = sep_1
        ) |>
        pivot_longer(
            cols = -id,
            names_to = c(".value", "pseudo_id"),
            names_pattern = "^(.+)_([1-3])$"
        )

    # Inclusion criteria:
    df <- df |>
        # (Pseudo-)participant is at risk of outcome.
        filter(chdout_bl == 0) |>
        # And reported the following baseline variables (all mediators included).
        drop_na(sex, age, sep, ends_with("bl"))

    df
}

impute_dataset <- function(df) {
```

```r
    exp_covars <- c("age", "sex", "sep")
    med_covars <- c("age", "sex", "sep",
                    "livingalone", "raremeets", "support01", "civstat01")

    impdf <- df |>
        mutate(sep = factor(sep)) |>
        # Exposures.
        impute_logreg("livingalone", exp_covars) |>
        impute_logreg("raremeets", exp_covars) |>
        impute_logreg("support01", exp_covars) |>
        impute_logreg("civstat01", exp_covars) |>
        # Metabolic mediators.
        impute_logreg("hbp", c("hbp_bl", med_covars)) |>
        impute_logreg("hc", c("hc_bl", med_covars)) |>
        impute_logreg("diab", c("diab_bl", med_covars)) |>
        impute_logreg("bmi01", c("bmi01_bl", med_covars)) |>
        # Behavioral mediators.
        impute_logreg("alco01", c("alco01_bl", med_covars)) |>
        impute_logreg("smoker", c("smoker_bl", med_covars)) |>
        impute_logreg("met01", c("met01_bl", med_covars)) |>
        impute_logreg("fruveg01", c("fruveg01_bl", med_covars)) |>
        impute_logreg("sleep01", c("sleep01_bl", med_covars)) |>
        # Composites.
        impute_logreg("metabmulti01", c("metabmulti01_bl", med_covars)) |>
        impute_logreg("behavmulti01", c("behavmulti01_bl", med_covars)) |>
        impute_logreg("polyrisk01", c("polyrisk01_bl", med_covars)) |>
        mutate(sep = as.integer(sep))

    impdf |>
        mutate(
            age2 = age^2,
            # Standardize age variables to help inference.
            age = (age - mean(age)) / (2*sd(age)),
            age2 = (age2 - mean(age2)) / (2*sd(age2)),
            # Socioeconomic position modelled using indicator variables
            # (standard although not ideal).
            sep2 = if_else(sep == 2, 1, 0),
            sep3 = if_else(sep == 3, 1, 0),
            sep4 = if_else(sep == 4, 1, 0),
            time = as.integer(pseudo_id)
        ) |>
        select(-pseudo_id, -sep)
}

# Function for calculating counts and %s with privacy and
# missingness reporting included.
count_perc <- function(x, remove_na = TRUE) {
    stopifnot(is.logical(x))
    n <- sum(x, na.rm = TRUE)

    if (remove_na) {
        N <- sum(!is.na(x))
        message(
            paste0(
                "Missing values removed: ",
```

```
                sum(is.na(x)),
                " (",
                round(100 * (sum(is.na(x)) / length(x)), digits = 0),
                " %)"
            )
        )
    } else {
        N <- length(x)
    }

    perc <- format(round(n/N*100, digits = 1), nsmall = 1)

    # 0 < n < 10 cells are removed for privacy.
    if (n != 0 & n < 10) {
        return("--")
    } else {
        return(paste0(n, " (", perc, ")"))
    }
}

summarize_baseline <- function(df, by) {
    df |>
    select(
        id, {{by}},
        sex, age, sep,
        livingalone, raremeets, civstat01, support01,
        hbp, hc, diab, bmi01,
        met01, smoker, alco01, fruveg01, sleep01
    ) |>
    group_by({{by}}) |>
    summarize(
        N = n(),
        # Confounders.
        `Female, # (%)` = count_perc(sex == 0),
        `(1) Years, mean` = format(round(mean(age, na.rm = TRUE), digits = 1), nsmall = 1),
        `(1) Manual, # (%)` = count_perc(sep == 4),
        `(1) Routine non-manual, # (%)` = count_perc(sep == 3),
        `(1) Semi-professional, # (%)` = count_perc(sep == 2),
        `(1) Manager/professional, # (%)` = count_perc(sep == 11),
        # Exposures.
        `(1) Yes, # (%)` = count_perc(livingalone == 1),
        `(1) < 2 meetups/month, # (%)` = count_perc(raremeets == 1),
        `(1) Score 0-4, # (%)` = count_perc(support01 == 1),
        `(1) Single, # (%)` = count_perc(civstat01 == 1),
        # Mediators.
        `(1) Has history, # (%)1` = count_perc(hbp == 1),
        `(1) Has history, # (%)2` = count_perc(hc == 1),
        `(1) Has history, # (%)3` = count_perc(diab == 1),
        `(1) Yes, # (%)2` = count_perc(bmi01 == 1),
        `(1) < 14 MET-hours/week, # (%)` = count_perc(met01 == 1),
        `(1) Current regular, # (%)` = count_perc(smoker == 1),
        `(1) > 7/14 units/week, # (%)` = count_perc(alco01 == 1),
        `(1) < 1 serving/day, # (%)` = count_perc(fruveg01 == 1),
        `(1) < 7 hours/workday, # (%)` = count_perc(sleep01 == 1)
    ) |>
    pivot_longer(cols = -subgroup, values_transform = as.character) |>
```

```
    pivot_wider(names_from = subgroup, values_from = value) |>
    mutate(name = str_remove(name, "[1-4]$")) |>
    mutate(
        variable = c("",
                     "Sex",
                     "Age",
                     rep("Occupational class", 4),
                     "Living alone",
                     "Friends/Relatives",
                     "Social support",
                     "Marital status",
                     "Hypertension",
                     "Hypercholesterolemia",
                     "Diabetes",
                     "Obesity (BMI 30+)",
                     "Physical activity",
                     "Smoking",
                     "Alcohol consumption",
                     "Fruit & vegetable consumption",
                     "Sleep duration")
    )
}

summarize_outcomes <- function(df) {
    df |>
        select(
            id, wave,
            subgroup,
            hbp, hc, diab, bmi01,
            met01, smoker, alco01, fruveg01, sleep01,
            ami, angpect, rose, chd_other
        ) |>
        pivot_wider(names_from = wave, values_from = subgroup:chd_other) |>
        mutate(
            new_hbp_2 = if_else(hbp_1 == 1, NA, hbp_2),
            new_diab_2 = if_else(diab_1 == 1, NA, diab_2),
            new_hc_2 = if_else(hc_1 == 1, NA, hc_2),
            # Calculate how many changed state.
            changed_bmi01_2 = as.integer(bmi01_1 != bmi01_2),
            changed_met01_2 = as.integer(met01_1 != met01_2),
            changed_smoker_2 = as.integer(smoker_1 != smoker_2),
            changed_alco01_2 = as.integer(alco01_1 != alco01_2),
            changed_fruveg01_2 = as.integer(fruveg01_1 != fruveg01_2),
            changed_sleep01_2 = as.integer(sleep01_1 != sleep01_2)
        ) |>
        mutate(
            # Same definition as main analysis outcome "chdout".
            cvd_345 = case_when(
                ami_3 == 1 | angpect_3 == 1 | rose_3 == 1 ~ 1L,
                chd_other_4 == 1 | rose_4 == 1 ~ 1L,
                chd_other_5 == 1 | rose_5 == 1 ~ 1L,
                chd_other_5 == 0 & rose_5 == 0 ~ 0L,
                TRUE ~ NA_integer_
            ),
            # Show proportion of Rose-only outcomes out of all.
            rose_345 = case_when(
```

```
                (rose_3 == 1 | rose_4 == 1 | rose_5 == 1) &
                    # Other items are 0 or missing so it's "Rose-only CVD".
                    (!(ami_3 %in% 1) &
                     !(angpect_3 %in% 1) &
                     !(chd_other_4 %in% 1) &
                     !(chd_other_5 %in% 1)) ~ 1L,
                cvd_345 == 1 | cvd_345 == 0 ~ 0L,
                TRUE ~ NA_integer_
            ),
            was_lost = is.na(cvd_345)
        ) |>
        group_by(subgroup_1) |>
        summarize(
            # Don't exclude NAs for outcome summaries (high missingness).
            `(3-5) Yes, # (%)1` = count_perc(cvd_345 == 1, remove_na = FALSE),
            `(3-5) Yes, # (%)2` = count_perc(rose_345 == 1, remove_na = FALSE),
            `(3-5) Yes, # (%)3` = count_perc(was_lost),
            `(2) New, # (%)1` = count_perc(new_hbp_2 == 1),
            `(2) New, # (%)2` = count_perc(new_hc_2 == 1),
            `(2) New, # (%)3` = count_perc(new_diab_2 == 1),
            `(2) Changed, # (%)1` = count_perc(changed_bmi01_2 == 1),
            `(2) Changed, # (%)2` = count_perc(changed_met01_2 == 1),
            `(2) Changed, # (%)3` = count_perc(changed_smoker_2 == 1),
            `(2) Changed, # (%)4` = count_perc(changed_alco01_2 == 1),
            `(2) Changed, # (%)5` = count_perc(changed_fruveg01_2 == 1),
            `(2) Changed, # (%)6` = count_perc(changed_sleep01_2 == 1)
        ) |>
        pivot_longer(cols = -subgroup_1) |>
        pivot_wider(names_from = subgroup_1, values_from = value) |>
        mutate(
            name = str_remove(name, "[1-9]$"),
            variable = c("Cardiovascular disease",
                         "Rose questionnaire only",
                         "Lost to follow-up",
                         "Hypertension",
                         "Hypercholesterolemia",
                         "Diabetes",
                         "Obesity (BMI 30+)",
                         "Physical activity",
                         "Smoking",
                         "Alcohol consumption",
                         "Fruit & vegetable consumption",
                         "Sleep duration")
        )
}

# Function for making Table 1.
tabulate_summaries_subset <- function(hhs, incl_ids) {

    # --- Phase 1 ---
    # Included column.
    tbl1_included <- hhs |>
        filter(wave == 1) |>
        mutate(subgroup = if_else(id %in% incl_ids, "Included", NA)) |>
        filter(subgroup == "Included") |>
        summarize_baseline(by = subgroup) |>
```

```r
    select(variable, name, Included)

# High, moderate, low social isolation, plus excluded columns.
tbl1_other <- hhs |>
    filter(wave == 1) |>
    mutate(included = id %in% incl_ids) |>
    rowwise() |>
    mutate(
        n_exposures = sum(
            c_across(c(livingalone, raremeets, support01, civstat01)),
            na.rm = FALSE
        )
    ) |>
    ungroup() |>
    mutate(
        subgroup = case_when(
            !included ~ "Excluded",
            n_exposures %in% 3:4 ~ "High (3-4)",
            n_exposures %in% 1:2 ~ "Moderate (1-2)",
            n_exposures == 0 ~ "Low (0)",
            TRUE ~ NA
        )
    ) |>
    summarize_baseline(by = subgroup) |>
    select(variable, name, `High (3-4)`, `Moderate (1-2)`, `Low (0)`, Excluded)

tbl1 <- tbl1_included |>
    left_join(tbl1_other, by = c("variable", "name"))

# --- Phases 2-5 ---
tbl2_included <- hhs |>
    mutate(subgroup = if_else(id %in% incl_ids, "Included", NA)) |>
    filter(subgroup == "Included") |>
    summarize_outcomes() |>
    # Due to journal limitations.
    filter(str_detect(name, "Changed|New", negate = TRUE)) |>
    select(variable, name, Included)

# High, moderate, low social isolation, plus excluded columns.
tbl2_other <- hhs |>
    mutate(included = id %in% incl_ids) |>
    rowwise() |>
    mutate(
        n_exposures = sum(
            c_across(c(livingalone, raremeets, support01, civstat01)),
            na.rm = FALSE
        )
    ) |>
    ungroup() |>
    mutate(
        subgroup = case_when(
            !included ~ "Excluded",
            n_exposures %in% 3:4 ~ "High (3-4)",
            n_exposures %in% 1:2 ~ "Moderate (1-2)",
            n_exposures == 0 ~ "Low (0)",
            TRUE ~ NA
```

```
            )
        ) |>
        summarize_outcomes() |>
        filter(str_detect(name, "Changed|New", negate = TRUE)) |>
        select(variable, name, `High (3-4)`, `Moderate (1-2)`, `Low (0)`, Excluded)

    tbl2 <- tbl2_included |>
        left_join(tbl2_other, by = c("variable", "name"))

    # Combine subtables and use flextable to render for hand-tuning.
    tbl1 |>
        bind_rows(tbl2) |>
        select(variable, name, Included, `High (3-4)`, `Moderate (1-2)`, `Low (0)`, Excluded) |>
        mutate(
            vargroup = c(
                "Total",
                rep("Background", 6),
                rep("Exposures: Social isolation", 4),
                rep("Mediators: Metabolic factors", 4),
                rep("Mediators: Health behaviours", 5),
                rep("Outcomes: CVD", 3)
            ),
            vargroup = factor(
                vargroup,
                levels = c("Total", "Background", "Exposures: Social isolation",
                           "Mediators: Metabolic factors", "Mediators: Health behaviours",
                           "Outcomes: CVD")
            )
        ) |>
        arrange(vargroup, variable) |>
        as_grouped_data(groups = "vargroup") |>
        as_flextable(hide_grouplabel = TRUE) |>
            theme_zebra() |>
            width(width = c(1, 1, rep(1, 1+3+1))) |>
            add_header_row(
                values = c("Variable", "Summary", "", "Social isolation*", ""),
                colwidths = c(1, 1, 1, 3, 1)
            ) |>
            set_header_labels(name = "", variable = "") |>
            merge_v(j = 2) |>
            align(j = 4:5, align = "center", part = "header") |>
            colformat_double(i = 2, j = 3:7, digits = 0) |>
            bold(i = ~ !is.na(vargroup)) |>
            bg(j = 1:2, bg = "white") |>
            hline(j = 1, i = c(4,8)) |>
            add_footer_lines(values = c(
                "* The number of exposures (0-4) in Phase 1.",
                "** Percentages calculated without missing values except for outcomes."
            ))
}

# Function for creating supplementary table S1.
tabulate_summaries_full <- function(hhs, incl_ids) {

    # --- Phase 1 ---
    # "Included" column.
```

```r
tbl1_included <- hhs |>
    filter(wave == 1) |>
    mutate(subgroup = if_else(id %in% incl_ids, "Included", "Excluded")) |>
    summarize_baseline(by = subgroup) |>
    select(variable, name, Included, Excluded)

# Two columns for each exposure.
tbl1_livingalone <- hhs |>
    filter(wave == 1) |>
    mutate(subgroup = if_else(livingalone == 1, "Yes", "No")) |>
    summarize_baseline(by = subgroup) |>
    select(variable, name, Yes, No)

tbl1_raremeets <- hhs |>
    filter(wave == 1) |>
    mutate(subgroup = if_else(raremeets == 1, "Rare", "Often")) |>
    summarize_baseline(by = subgroup) |>
    select(variable, name, Rare, Often)

tbl1_civstat01 <- hhs |>
    filter(wave == 1) |>
    mutate(subgroup = if_else(civstat01 == 1, "Singles", "Couples")) |>
    summarize_baseline(by = subgroup) |>
    select(variable, name, Singles, Couples)

tbl1_support01 <- hhs |>
    filter(wave == 1) |>
    mutate(subgroup = if_else(support01 == 1, "0-4 points", "5+ points")) |>
    summarize_baseline(by = subgroup) |>
    select(variable, name, `0-4 points`, `5+ points`)

tbl1 <- tbl1_included |>
    left_join(tbl1_livingalone, by = c("variable", "name")) |>
    left_join(tbl1_raremeets, by = c("variable", "name")) |>
    left_join(tbl1_civstat01, by = c("variable", "name")) |>
    left_join(tbl1_support01, by = c("variable", "name"))

# --- Phases 2-5 ---
tbl2_included <- hhs |>
    mutate(subgroup = if_else(id %in% incl_ids, "Included", "Excluded")) |>
    summarize_outcomes() |>
    select(variable, name, Included, Excluded)

# Two columns for each exposure.
tbl2_livingalone <- hhs |>
    mutate(subgroup = if_else(livingalone == 1, "Yes", "No")) |>
    summarize_outcomes() |>
    select(variable, name, Yes, No)

tbl2_raremeets <- hhs |>
    mutate(subgroup = if_else(raremeets == 1, "Rare", "Often")) |>
    summarize_outcomes() |>
    select(variable, name, Rare, Often)

tbl2_civstat01 <- hhs |>
    mutate(subgroup = if_else(civstat01 == 1, "Singles", "Couples")) |>
```

```
        summarize_outcomes() |>
        select(variable, name, Singles, Couples)

    tbl2_support01 <- hhs |>
        mutate(subgroup = if_else(support01 == 1, "0-4 points", "5+ points")) |>
        summarize_outcomes() |>
        select(variable, name, `0-4 points`, `5+ points`)

    tbl2 <- tbl2_included |>
        left_join(tbl2_livingalone, by = c("variable", "name")) |>
        left_join(tbl2_raremeets, by = c("variable", "name")) |>
        left_join(tbl2_civstat01, by = c("variable", "name")) |>
        left_join(tbl2_support01, by = c("variable", "name"))

    # Combine subtables and use flextable to visualize.
    tbl1 |>
        bind_rows(tbl2) |>
        select(variable, name, Included, Yes:`5+ points`, Excluded) |>
        mutate(
            vargroup = c(
                "Total",
                rep("Background", 6),
                rep("Exposures: Social isolation", 4),
                rep("Mediators: Metabolic factors", 4),
                rep("Mediators: Health behaviours", 5),
                rep("Outcomes: CVD", 3),
                rep("Mediators: Metabolic factors", 4),
                rep("Mediators: Health behaviours", 5)
            ),
            vargroup = factor(
                vargroup,
                levels = c("Total", "Background", "Exposures: Social isolation",
                           "Mediators: Metabolic factors", "Mediators: Health behaviours",
                           "Outcomes: CVD")
            )
        ) |>
        arrange(vargroup, variable) |>
        as_grouped_data(groups = "vargroup") |>
        as_flextable(hide_grouplabel = TRUE) |>
            theme_zebra() |>
            width(width = c(2, 2.7, rep(1.25, 1+8+1))) |>
            add_header_row(
                values = c("Variable", "Summary", "",
                           "Living alone", "Meeting\nfriends/relatives",
                           "Marital status", "Social support", ""),
                colwidths = c(1, 1, 1, 2, 2, 2, 2, 1)
            ) |>
            set_header_labels(name = "", variable = "") |>
            merge_v(j = 2) |>
            align(j = 4:11, align = "center", part = "header") |>
            colformat_double(i = 2, j = 3:7, digits = 0) |>
            bold(i = ~ !is.na(vargroup)) |>
            bg(bg = "white", part = "all") |>
            hline(j = 1:12, i = c(4,8,17,19,21,26,28,30,32)) |>
            vline(j = c(3,5,7,9,11), part = "body")
}
```

```
# Total effect Stan model without mediation.
bayesgtotal <- function(df, A, Y, L) {
    stanmodel <- '
        data {
          int N;
          int N_obs;
          array[N_obs] int idx_obs;

          int P_L;
          int P_A;
          int N_T;

          matrix[N, P_L] L; // Confounders.
          matrix[N, P_A] A; // Exposures.
          array[N] int Y; // Outcome.
          array[N] int time;

          // Interventions of interest.
          matrix[N,P_A] A1111;
          matrix[N,P_A] A0000;
          matrix[N,P_A] A1ooo;
          matrix[N,P_A] A0ooo;
          matrix[N,P_A] Ao1oo;
          matrix[N,P_A] Ao0oo;
          matrix[N,P_A] Aoo1o;
          matrix[N,P_A] Aoo0o;
          matrix[N,P_A] Aooo1;
          matrix[N,P_A] Aooo0;
        }

        transformed data {
          matrix[N_obs, P_L] L_obs = L[idx_obs,:];
          matrix[N_obs, P_A] A_obs = A[idx_obs,:];
          array[N_obs] int Y_obs = Y[idx_obs];
          array[N_obs] int time_obs = time[idx_obs];
        }

        parameters {
          vector[P_L] alphaL;
          vector[P_A] alphaA;
          vector[N_T] alphaT;
          real alpha0;
        }

        model {
          alpha0 ~ normal(0, 5);
          alphaT ~ normal(0, 2);
          alphaA ~ normal(0, 2);
          alphaL ~ normal(0, 2);

          Y_obs ~ bernoulli_logit(
            alpha0 + alphaT[time_obs] + A_obs*alphaA + L_obs*alphaL
          );
        }
```

```
    generated quantities {
      real RR_0; // 0 refers to A1111 / A0000
      real RR_1; // 1 refers to A1ooo / A0ooo
      real RR_2; // ...
      real RR_3;
      real RR_4;

      {
          vector[N] etaY = alpha0 + alphaT[time] + L*alphaL;

          real EPrYa1_0 = mean(inv_logit(etaY + A1111*alphaA));
          real EPrYa0_0 = mean(inv_logit(etaY + A0000*alphaA));

          real EPrYa1_1 = mean(inv_logit(etaY + A1ooo*alphaA));
          real EPrYa0_1 = mean(inv_logit(etaY + A0ooo*alphaA));

          real EPrYa1_2 = mean(inv_logit(etaY + Ao1oo*alphaA));
          real EPrYa0_2 = mean(inv_logit(etaY + Ao0oo*alphaA));

          real EPrYa1_3 = mean(inv_logit(etaY + Aoo1o*alphaA));
          real EPrYa0_3 = mean(inv_logit(etaY + Aoo0o*alphaA));

          real EPrYa1_4 = mean(inv_logit(etaY + Aooo1*alphaA));
          real EPrYa0_4 = mean(inv_logit(etaY + Aooo0*alphaA));

          RR_0 = EPrYa1_0 / EPrYa0_0;
          RR_1 = EPrYa1_1 / EPrYa0_1;
          RR_2 = EPrYa1_2 / EPrYa0_2;
          RR_3 = EPrYa1_3 / EPrYa0_3;
          RR_4 = EPrYa1_4 / EPrYa0_4;
      }
    }

'

df_all <- df |> as.data.frame()
df_all[is.na(df_all[[Y]]), Y] <- 999 # Stan doesn't like NAs.

# Interventions of interest.
A1111 <- matrix(1, nrow(df_all), length(A))
A0000 <- matrix(0, nrow(df_all), length(A))
A1ooo <- as.matrix(df_all[, A]); A1ooo[, 1] <- 1
A0ooo <- as.matrix(df_all[, A]); A0ooo[, 1] <- 0
Ao1oo <- as.matrix(df_all[, A]); Ao1oo[, 2] <- 1
Ao0oo <- as.matrix(df_all[, A]); Ao0oo[, 2] <- 0
Aoo1o <- as.matrix(df_all[, A]); Aoo1o[, 3] <- 1
Aoo0o <- as.matrix(df_all[, A]); Aoo0o[, 3] <- 0
Aooo1 <- as.matrix(df_all[, A]); Aooo1[, 4] <- 1
Aooo0 <- as.matrix(df_all[, A]); Aooo0[, 4] <- 0

res <- rstan::stan(
    model_code = stanmodel,
    iter = 3000,
    warmup = 1000,
    data = list(
        # Dimensions.
```

```
            N = nrow(df_all), # Number of pseudo-participants.
            N_obs = which(df_all[[Y]] != 999) |> length(), # Number non-censored.
            P_L = length(L),
            P_A = length(A),
            N_T = length(unique(df_all$time)), # Number of time points (3).
            idx_obs = which(df_all[[Y]] != 999),
            # Data.
            time = df_all$time,
            Y = df_all[[Y]],
            A = as.matrix(df_all[, A]),
            L = as.matrix(df_all[, L]),
            # Interventions.
            A1111 = A1111,
            A0000 = A0000,
            A1ooo = A1ooo,
            A0ooo = A0ooo,
            Ao1oo = Ao1oo,
            Ao0oo = Ao0oo,
            Aoo1o = Aoo1o,
            Aoo0o = Aoo0o,
            Aooo1 = Aooo1,
            Aooo0 = Aooo0
        )
    )

    res |>
        rstan::summary(
            pars = c("RR_0", "RR_1", "RR_2", "RR_3", "RR_4"),
            probs = c(0.05, 0.95) # 90% credible interval via quantiles.
        ) |>
        pluck("summary") |>
        as_tibble(rownames = "parameter") |>
        select(parameter, mean, ci05 = `5%`, ci95 = `95%`) |>
        pivot_wider(
            names_from = parameter,
            values_from = c(ci05, ci95, mean)
        ) |>
        mutate(Y = Y) |>
        select(Y, everything())
}

# Multi-exposure multi-mediator model. (Single-mediator model is a special case.)
bayesgmed <- function(df, A, M, Y, L) {
    stanmodel <- '
        data {
          int<lower=0> N; // Number of pseudo-participants.
          int<lower=0> N_obs;
          array[N_obs] int idx_obs; // Indeces with no missing outcomes.
          int<lower=0> P_L;
          int<lower=0> P_A;
          int<lower=0> P_M;
          int N_T;

          array[N] int time;
          matrix[N, P_A] A;
          array[N, P_M] int M; // Mediators.
```

```
  array[N] int Y;
  matrix[N, P_L] L;

  // Interventions of interest.
  matrix[N,P_A] A1111;
  matrix[N,P_A] A0000;
  matrix[N,P_A] A1ooo;
  matrix[N,P_A] A0ooo;
  matrix[N,P_A] Ao1oo;
  matrix[N,P_A] Ao0oo;
  matrix[N,P_A] Aoo1o;
  matrix[N,P_A] Aoo0o;
  matrix[N,P_A] Aooo1;
  matrix[N,P_A] Aooo0;
}

transformed data {
  matrix[N_obs, P_L] L_obs = L[idx_obs,:];
  matrix[N_obs, P_A] A_obs = A[idx_obs,:];
  matrix[N_obs, P_M] Mm_obs = to_matrix(M[idx_obs,:]);
  array[N_obs, P_M] int M_obs = M[idx_obs,:];
  array[N_obs] int Y_obs = Y[idx_obs];
  array[N_obs] int time_obs = time[idx_obs];
}

parameters {
  // Outcome model (labelled alpha).
  vector[P_L] alphaL;
  vector[P_M] alphaM;
  vector[P_A] alphaA;
  vector[N_T] alphaT;
  real alpha0;

  // Mediator models (labelled beta).
  array[P_M] vector[P_L] betaL;
  array[P_M] vector[P_A] betaA;
  array[P_M] vector[N_T] betaT;
  vector[P_M] beta0;
}

model {
  alpha0 ~ normal(0, 5);
  alphaA ~ normal(0, 2);
  alphaL ~ normal(0, 2);
  alphaM ~ normal(0, 2);
  alphaT ~ normal(0, 2);

 for (p in 1:P_M) {
      beta0 ~ normal(0, 5);
      betaL[p] ~ normal(0, 2);
      betaA[p] ~ normal(0, 2);
      betaT[p] ~ normal(0, 2);
  }

  for (p in 1:P_M) {
      M_obs[:,p] ~ bernoulli_logit(
```

```
        beta0[p] + betaT[p][time_obs] + A_obs*betaA[p] + L_obs*betaL[p]
      );
    }

    Y_obs ~ bernoulli_logit(
      alpha0 + alphaT[time_obs] + A_obs*alphaA + Mm_obs*alphaM + L_obs*alphaL
    );
  }

  generated quantities {
    real NDE_RR_0; // Natural direct effect.
    real NDE_RR_1;
    real NDE_RR_2;
    real NDE_RR_3;
    real NDE_RR_4;

    real NIE_RR_0; // Natural indirect effect.
    real NIE_RR_1;
    real NIE_RR_2;
    real NIE_RR_3;
    real NIE_RR_4;
    {
        // Simulate mediators under each intervention.
        matrix[N,P_M] Ma1_0;
        matrix[N,P_M] Ma0_0;
        matrix[N,P_M] Ma1_1;
        matrix[N,P_M] Ma0_1;
        matrix[N,P_M] Ma1_2;
        matrix[N,P_M] Ma0_2;
        matrix[N,P_M] Ma1_3;
        matrix[N,P_M] Ma0_3;
        matrix[N,P_M] Ma1_4;
        matrix[N,P_M] Ma0_4;

        vector[N] etaM;

        for (p in 1:P_M) {
          etaM = beta0[p] + betaT[p][time] + L*betaL[p];

          Ma1_0[:,p] = to_vector(bernoulli_logit_rng(etaM + A1111*betaA[p]));
          Ma0_0[:,p] = to_vector(bernoulli_logit_rng(etaM + A0000*betaA[p]));

          Ma1_1[:,p] = to_vector(bernoulli_logit_rng(etaM + A1ooo*betaA[p]));
          Ma0_1[:,p] = to_vector(bernoulli_logit_rng(etaM + A0ooo*betaA[p]));

          Ma1_2[:,p] = to_vector(bernoulli_logit_rng(etaM + Ao1oo*betaA[p]));
          Ma0_2[:,p] = to_vector(bernoulli_logit_rng(etaM + Ao0oo*betaA[p]));

          Ma1_3[:,p] = to_vector(bernoulli_logit_rng(etaM + Aoo1o*betaA[p]));
          Ma0_3[:,p] = to_vector(bernoulli_logit_rng(etaM + Aoo0o*betaA[p]));

          Ma1_4[:,p] = to_vector(bernoulli_logit_rng(etaM + Aooo1*betaA[p]));
          Ma0_4[:,p] = to_vector(bernoulli_logit_rng(etaM + Aooo0*betaA[p]));
        }

        // Pre-compute common terms...
```

```
            vector[N] etaY = alpha0 + alphaT[time] + L*alphaL;

            vector[N] etaY_a1_0 = etaY + A1111*alphaA;
            vector[N] etaY_a1_1 = etaY + A1ooo*alphaA;
            vector[N] etaY_a1_2 = etaY + Ao1oo*alphaA;
            vector[N] etaY_a1_3 = etaY + Aoo1o*alphaA;
            vector[N] etaY_a1_4 = etaY + Aooo1*alphaA;

            // Simulate outcomes under interventions and simulated mediators.
            // For example, "EPrYa1ma1_0" can be read as "expected probability of outcome
            // under a=1 and M under a=1, where a=1 is the intervention A1111 (all 1)".
            real EPrYa1ma1_0 = mean(inv_logit(etaY_a1_0 + Ma1_0*alphaM));
            real EPrYa1ma0_0 = mean(inv_logit(etaY_a1_0 + Ma0_0*alphaM));
            real EPrYa0ma0_0 = mean(inv_logit(etaY + A0000*alphaA + Ma0_0*alphaM));

            real EPrYa1ma1_1 = mean(inv_logit(etaY_a1_1 + Ma1_1*alphaM));
            real EPrYa1ma0_1 = mean(inv_logit(etaY_a1_1 + Ma0_1*alphaM));
            real EPrYa0ma0_1 = mean(inv_logit(etaY + A0ooo*alphaA + Ma0_1*alphaM));

            real EPrYa1ma1_2 = mean(inv_logit(etaY_a1_2 + Ma1_2*alphaM));
            real EPrYa1ma0_2 = mean(inv_logit(etaY_a1_2 + Ma0_2*alphaM));
            real EPrYa0ma0_2 = mean(inv_logit(etaY + Ao0oo*alphaA + Ma0_2*alphaM));

            real EPrYa1ma1_3 = mean(inv_logit(etaY_a1_3 + Ma1_3*alphaM));
            real EPrYa1ma0_3 = mean(inv_logit(etaY_a1_3 + Ma0_3*alphaM));
            real EPrYa0ma0_3 = mean(inv_logit(etaY + Aoo0o*alphaA + Ma0_3*alphaM));

            real EPrYa1ma1_4 = mean(inv_logit(etaY_a1_4 + Ma1_4*alphaM));
            real EPrYa1ma0_4 = mean(inv_logit(etaY_a1_4 + Ma0_4*alphaM));
            real EPrYa0ma0_4 = mean(inv_logit(etaY + Aooo0*alphaA + Ma0_4*alphaM));

            // NDE is the effect of a=1 vs. a=0 holding M constant under a=0.
            NDE_RR_0 = EPrYa1ma0_0 / EPrYa0ma0_0;
            NDE_RR_1 = EPrYa1ma0_1 / EPrYa0ma0_1;
            NDE_RR_2 = EPrYa1ma0_2 / EPrYa0ma0_2;
            NDE_RR_3 = EPrYa1ma0_3 / EPrYa0ma0_3;
            NDE_RR_4 = EPrYa1ma0_4 / EPrYa0ma0_4;

            // NIE is the effect of M under a=1 vs. a=0 holding A constant at a=1.
            NIE_RR_0 = EPrYa1ma1_0 / EPrYa1ma0_0;
            NIE_RR_1 = EPrYa1ma1_1 / EPrYa1ma0_1;
            NIE_RR_2 = EPrYa1ma1_2 / EPrYa1ma0_2;
            NIE_RR_3 = EPrYa1ma1_3 / EPrYa1ma0_3;
            NIE_RR_4 = EPrYa1ma1_4 / EPrYa1ma0_4;
          }
      }
  '
  df_all <- df |> as.data.frame()
  df_all[is.na(df_all[[Y]]), Y] <- 999 # Stan doesn't like NAs.

  # Interventions of interest.
  A1111 <- matrix(1, nrow(df_all), length(A))
  A0000 <- matrix(0, nrow(df_all), length(A))
  A1ooo <- as.matrix(df_all[, A]); A1ooo[, 1] <- 1
  A0ooo <- as.matrix(df_all[, A]); A0ooo[, 1] <- 0
  Ao1oo <- as.matrix(df_all[, A]); Ao1oo[, 2] <- 1
```

```
    Ao0oo <- as.matrix(df_all[, A]); Ao0oo[, 2] <- 0
    Aoo1o <- as.matrix(df_all[, A]); Aoo1o[, 3] <- 1
    Aoo0o <- as.matrix(df_all[, A]); Aoo0o[, 3] <- 0
    Aooo1 <- as.matrix(df_all[, A]); Aooo1[, 4] <- 1
    Aooo0 <- as.matrix(df_all[, A]); Aooo0[, 4] <- 0

    res <- rstan::stan(
        model_code = stanmodel,
        iter = 3000, # 2000 * 4 (# of cores) = 8000 samples drawn in total.
        warmup = 1000,
        data = list(
            # Dimensions.
            P_L = length(L),
            P_A = length(A),
            P_M = length(M),
            N_T = length(unique(df_all$time)),
            N = nrow(df_all),
            N_obs = length(which(df_all[[Y]] != 999)),
            idx_obs = which(df_all[[Y]] != 999),
            # Data.
            time = df_all$time,
            Y = df_all[[Y]],
            A = as.matrix(df_all[, A]),
            M = as.matrix(df_all[, M]),
            L = as.matrix(df_all[, L]),
            # Interventions.
            A1111 = A1111,
            A0000 = A0000,
            A1ooo = A1ooo,
            A0ooo = A0ooo,
            Ao1oo = Ao1oo,
            Ao0oo = Ao0oo,
            Aoo1o = Aoo1o,
            Aoo0o = Aoo0o,
            Aooo1 = Aooo1,
            Aooo0 = Aooo0
        )
    )

    res |>
        rstan::summary(
            pars = c("NDE_RR_0", "NDE_RR_1", "NDE_RR_2", "NDE_RR_3", "NDE_RR_4",
                     "NIE_RR_0", "NIE_RR_1", "NIE_RR_2", "NIE_RR_3", "NIE_RR_4"),
            probs = c(0.05, 0.95)
        ) |>
        pluck("summary") |>
        as_tibble(rownames = "parameter") |>
        select(parameter, mean, ci05 = `5%`, ci95 = `95%`) |>
        pivot_wider(
            names_from = parameter,
            values_from = c(ci05, ci95, mean)
        ) |>
        mutate(M = list(M), Y = Y) |>
        select(M, Y, everything())
}
```

```
# Function to run total effects model for each outcome.
run_totaleffects <- function(hhs_incl, vars) {
    expand_grid(
            A = list(vars[["A"]]),
            Y = c(vars[["Y"]], vars[["M2"]]),
            L = list(vars[["L"]])
        ) |>
        # If outcome is mediator, add baseline value to confounders L.
        # Baseline CVD is handled by exclusion (not at risk).
        rowwise() |>
        mutate(L = if_else(Y == "chdout", list(L), list(c(paste0(Y,"_bl"), L)))) |>
        ungroup() |>
        pmap(
            \(A,Y,L) bayesgtotal(hhs_incl, A, Y, L),
            .progress = "Total effect analyses"
        ) |>
        list_rbind()
}

# Function to run mediation analysis for each mediator separately.
run_single_medeffects <- function(hhs_incl, vars) {
    expand_grid(
            A = list(vars[["A"]]),
            M = vars[["M2"]],
            Y = vars[["Y"]],
            L = list(vars[["L"]])
        ) |>
        # Add baseline mediator to confounders L.
        rowwise() |>
        mutate(L = list(c(paste0(M,"_bl"), L))) |>
        ungroup() |>
        pmap(
            \(A,M,Y,L) bayesgmed(hhs_incl, A, M, Y, L),
            .progress = "Single-mediator analyses"
        ) |>
        list_rbind()
}

# Table 2. Total effect analysis results.
tabulate_totresults <- function(x) {

    x |>
        select(
            Y,
            mean_RR_0, ci05_RR_0, ci95_RR_0,
            mean_RR_1, ci05_RR_1, ci95_RR_1,
            mean_RR_2, ci05_RR_2, ci95_RR_2,
            mean_RR_3, ci05_RR_3, ci95_RR_3,
            mean_RR_4, ci05_RR_4, ci95_RR_4
        ) |>
        pivot_longer(cols = mean_RR_0:ci95_RR_4) |>
        separate(col = name, into = c("variable", "A"), sep = -2) |>
        mutate(value = format(round(value, digits = 2), nsmall = 2)) |>
        pivot_wider(names_from = variable, values_from = "value") |>
        mutate(RR = str_c(mean_RR," (",ci05_RR,"-",ci95_RR,")")) |>
        select(-(mean_RR:ci95_RR)) |>
```

```r
        pivot_wider(names_from = A, values_from = RR) |>
        mutate(
            Y = factor(
                Y,
                levels = c("chdout", "hbp", "diab", "hc", "bmi01",
                           "smoker", "alco01", "sleep01", "fruveg01",
                           "met01", "metabmulti01", "behavmulti01",
                           "polyrisk01"),
                labels = c("Cardiovascular disease",
                           "Hypertension",
                           "Diabetes",
                           "Hypercholesterolemia",
                           "Obesity (BMI 30+)",
                           "Smoking",
                           "Heavy alcohol consumption",
                           "Short sleep duration",
                           "Low fruit & vegetable\nconsumption",
                           "Physical inactivity",
                           "2+ metabolic factors",
                           "2+ behavioral factors",
                           "4+ risk factors")
            ),
            vargroup = c("",
                         rep("Metabolic factors", 4),
                         rep("Health behaviors", 5),
                         rep("Mediator composites", 3)
            )
        ) |>
        as_grouped_data(groups = "vargroup") |>
        as_flextable(hide_grouplabel = TRUE) |>
            theme_zebra() |>
            set_header_labels(
                values = c("",
                           "Social isolation\n(all four vs. none)",
                           "Living alone",
                           "Rare meetups\n(< twice in 4 weeks)",
                           "Low social support\n(0-4 points)",
                           "Single\n(marital status)")
            ) |>
            add_header_row(
                values = c("Outcome", "Exposure"),
                colwidths = c(1, 5)
            ) |>
            width(width = c(2.5, rep(1.6, 5))) |>
            align(j = 2:6, align = "center", part = "header") |>
            bold(i = ~ !is.na(vargroup)) |>
            bg(i = ~ !is.na(vargroup), bg = "white")
}

# Supplementary table S2. Mediation analysis results.
tabulate_medresults <- function(medresults) {

    medresults |>
        select(
            M,
            mean_NDE_RR_0, ci05_NDE_RR_0, ci95_NDE_RR_0,
```

```
        mean_NDE_RR_1, ci05_NDE_RR_1, ci95_NDE_RR_1,
        mean_NDE_RR_2, ci05_NDE_RR_2, ci95_NDE_RR_2,
        mean_NDE_RR_3, ci05_NDE_RR_3, ci95_NDE_RR_3,
        mean_NDE_RR_4, ci05_NDE_RR_4, ci95_NDE_RR_4,
        mean_NIE_RR_0, ci05_NIE_RR_0, ci95_NIE_RR_0,
        mean_NIE_RR_1, ci05_NIE_RR_1, ci95_NIE_RR_1,
        mean_NIE_RR_2, ci05_NIE_RR_2, ci95_NIE_RR_2,
        mean_NIE_RR_3, ci05_NIE_RR_3, ci95_NIE_RR_3,
        mean_NIE_RR_4, ci05_NIE_RR_4, ci95_NIE_RR_4
    ) |>
    pivot_longer(cols = mean_NDE_RR_0:ci95_NIE_RR_4) |>
    separate(col = name, into = c("variable", "A"), sep = -2) |>
    mutate(value = format(round(value, digits = 2), nsmall = 2)) |>
    mutate(
        effect = if_else(
            str_detect(variable, "NDE"),
            true = "Direct",
            false = "Indirect"
        ),
        variable = str_remove(variable, "_NDE|_NIE")
    ) |>
    pivot_wider(names_from = variable, values_from = "value") |>
    mutate(RR = str_c(mean_RR," (",ci05_RR,"-",ci95_RR,")")) |>
    select(-(mean_RR:ci95_RR)) |>
    pivot_wider(names_from = A, values_from = RR) |>
    mutate(
        M = factor(
            M,
            levels = c("hbp", "diab", "hc", "bmi01",
                       "smoker", "alco01", "sleep01", "fruveg01",
                       "met01", "metabmulti01", "behavmulti01",
                       "polyrisk01", "Metabolic factors", "Behavioral factors",
                       "All factors"),
            labels = c("Hypertension",
                       "Diabetes",
                       "Hypercholesterolemia",
                       "Obesity (BMI 30+)",
                       "Smoking",
                       "Heavy alcohol consumption",
                       "Short sleep\nduration",
                       "Low fruit & vegetable\nconsumption",
                       "Physical inactivity",
                       "2+ metabolic factors",
                       "2+ behavioral factors",
                       "4+ risk factors",
                       "Metabolic factors",
                       "Behavioral factors",
                       "All factors")
        ),
        vargroup = c(rep("Metabolic factors", 4*2),
                     rep("Health behaviors", 5*2),
                     rep("Mediator composites", 3*2),
                     rep("Multi-mediators models", 3*2)
        )
    ) |>
    as_grouped_data(groups = "vargroup") |>
```

```r
        as_flextable(hide_grouplabel = TRUE) |>
            theme_zebra() |>
            set_header_labels(
                values = c("",
                           "",
                           "Social isolation\n(all vs. none)",
                           "Living alone",
                           "Rare meetups\n(< 2/month)",
                           "Low support\n(0-4 points)",
                           "Single\n(marital status)")
            ) |>
            add_header_row(
                values = c("Mediator", "Pathway", "Exposure"),
                colwidths = c(1, 1, 5)
            ) |>
            width(width = c(2, 0.75, rep(1.5, 5))) |>
            align(i = 1, j = 3:7, align = "center", part = "header") |>
            bold(i = ~ !is.na(vargroup)) |>
            merge_v(j = 2) |>
            bg(bg = "white", part = "all") |>
            hline(i = c(3,5,7, 12,14,16,18, 23,25, 30,32))
}

run_analysis <- function() {
    rawdir <- "path/to/raw-data/"
    tidyfile <- "path/to/tidy-data.RData"
    resultfile <- "path/to/results.RData"

    import_tidydata(update = FALSE, tidyfile, rawdir, varnamefile = varname_str)

    hhs_incl_na <- prepare_dataset(hhs) # Study design.
    hhs_incl <- impute_dataset(hhs_incl_na) # Imputation.

    # Variables.
    vars <- list(
        L = c("sex", "sep2", "sep3", "sep4", "age", "age2"),
        A = c("livingalone",
              "raremeets",
              "support01",
              "civstat01"),
        M2 = c("hbp", "diab", "hc", "bmi01",
               "smoker", "alco01", "sleep01", "fruveg01", "met01",
               "metabmulti01", "behavmulti01", "polyrisk01"),
        Y = "chdout",
        M1 = c("hbp_bl", "diab_bl", "hc_bl", "bmi01_bl",
               "smoker_bl", "alco01_bl", "sleep01_bl", "fruveg01_bl", "met01_bl")
    )

    totresults <- run_totaleffects(hhs_incl, vars)
    singlemedresults <- run_single_medeffects(hhs_incl, vars)
    # Multi-mediator analyses.
    metabmedresults <- bayesgmed(
        df = hhs_incl,
        A = vars[["A"]],
        M = vars[["M2"]][1:4], # Metabolic risk factors.
        Y = vars[["Y"]],
```

```r
    L = c(vars[["L"]], vars[["M1"]][1:4])
)
gc()
behavmedresults <- bayesgmed(
    df = hhs_incl,
    A = vars[["A"]],
    M = vars[["M2"]][5:9], # Behavioural risk factors.
    Y = vars[["Y"]],
    L = c(vars[["L"]], vars[["M1"]][5:9])
)
gc()
allmedresults <- bayesgmed(
    df = hhs_incl,
    A = vars[["A"]],
    M = vars[["M2"]][1:9], # All studied mediators.
    Y = vars[["Y"]],
    L = c(vars[["L"]], vars[["M1"]][1:9])
)
gc()

results <<- list(
    total = totresults,
    singlemed = singlemedresults,
    behavmed = behavmedresults,
    metabmed = metabmedresults,
    allmed = allmedresults
)
save(results, file = resultfile)

# Basic statistics.
descriptive_stats <<- list(
    n_incl = length(unique(hhs_incl$id)), # Participants with at least one observation.
    prop_incl = n_incl / length(unique(hhs$id)),
    nt_incl = nrow(hhs_incl), # Observations.
    nt_incl_cens = length(which(is.na(hhs_incl$chdout))),
    prop_incl_cens = nt_incl_cens / nt_incl, # Censored observations.
)

# Table 1.
tbl1_subset <<- tabulate_summaries_subset(hhs, incl_ids = unique(hhs_incl_na$id))
save_as_docx(tbl1_subset, path = "path/to/manuscript/table1.docx")
save_as_image(tbl1_subset, path = "path/to/manuscript/table1.png")

# Supplementary table S1 for more descriptive statistics.
tbl1_full <<- tabulate_summaries_full(hhs, incl_ids = unique(hhs_incl_na$id))
save_as_image(tbl1_full, path = "path/to/manuscript/table1-full.png", res = 1000)

# Table 2 (total effect results).
tottable <<- tabulate_totresults(results$total)
save_as_image(tottable, path = "path/to/manuscript/table2.png")
save_as_docx(tottable, path = "path/to/manuscript/table2.docx")

# Supplementary table S2 (mediation analysis results).
multimedresults <- bind_rows(
    mutate(results$behavmed, M = "Behavioral factors"),
    mutate(results$metabmed, M = "Metabolic factors"),
```

```
        mutate(results$allmed, M = "All factors")
    )
    medtable <<- results$singlemed |>
        mutate(M = as.character(M)) |>
        bind_rows(multimedresults) |>
        tabulate_medresults()

    save_as_image(medtable, path = "path/to/manuscript/table3.png", res = 300)

    # Proportion isolated (all 1) by survey phase.
    prop_isol <<- hhs |>
        filter(id %in% hhs_incl_na$id) |>
        mutate(is_isol = livingalone == 1 & raremeets == 1 &
                         civstat01 == 1 & support01 == 1
        ) |>
        group_by(wave) |>
        summarize(
            n = n(),
            n_obs = sum(!is.na(is_isol)),
            n_isol = sum(is_isol, na.rm = TRUE),
            prop_isol = n_isol / n_obs
        )

    # Less than 2.5% missing in observation dataset.
    # Most in infrequent meetups (2.49%).
    prop_miss <<- hhs_incl_na |>
        summarize(across(everything(), \(x) sum(is.na(x)) / length(x))) |>
        pivot_longer(cols = everything()) |>
        arrange(value)

}

varname_str <- "
new_name,old_name
...
"
```
